# Nicotine-replacement therapy, as a surrogate of smoking, and the risk of hospitalization with Covid-19 and all-cause mortality: a nationwide, observational cohort study in France

**DOI:** 10.1101/2020.07.28.20160630

**Authors:** Mahmoud Zureik, Bérangère Baricault, Clémentine Vabre, Laura Semenzato, Jérôme Drouin, François Cuenot, Laetitia Penso, Philippe Herlemont, Emilie Sbidian, Alain Weill, Mathieu Molimard, Rosermary Dray-Spira, Jérémie Botton

## Abstract

**Introduction:** Several studies have reported an unexpectedly low prevalence of current smoking among hospitalized patients with Covid-19. However, these studies mostly compared observed to expected rates of smoking without direct comparison with individual controls.

**Objective:** To examine the association of nicotine-replacement therapy, as a surrogate of smoking, with hospitalization and all-cause mortality during the first wave of SARS-CoV-2 epidemic in France.

**Methods:** We conducted a nationwide matched “exposed/unexposed” cohort study using information from the French national health data system which covers the entire French population. We conducted two separate analyses, the first in individuals exposed to nicotine-replacement therapy without major smoking-related diseases (cancer, cardiovascular and/or respiratory diseases) and the second in those presenting these conditions. We included all individuals, aged between 18 and 75 years, who had been reimbursed at least one nicotine-replacement therapy between November 15, 2019, and February 15, 2020. For each exposed individual, we randomly selected, from the entire Metropolitan French population, up to two non-exposed individuals (1:2) matched for the following variables: age (same year of birth), sex, department of residence (n=96 in Metropolitan France), and complementary universal health insurance (CMU-C). The three end points were a hospitalization with Covid-19, a death or an intubation in hospitalized patients with Covid-19, and all-cause mortality. We compared outcomes in individuals who were exposed to nicotine-replacement therapy with those in individuals who were not, using a multivariable Cox model with inverse probability weighting according to the propensity score.

**Results:** In the first analysis, 297,070 individuals without major smoking-related diseases exposed to nicotine-replacement therapy were matched with 558,228 unexposed individuals without major smoking-related diseases. Individuals were aged on average 45.6 years (standard deviation: 12.7) and 48.8% were male.

From February 15, 2020 to June 7, 2020, hospitalization with Covid-19 occurred in 647 patients (151 patients in the nicotine-replacement therapy group and 496 patients in the unexposed group). In the main multivariable analysis, nicotine-replacement therapy was associated with a decreased risk of hospitalization with Covid-19 compared with unexposed individuals (hazard ratio, 0.50; 95% CI, 0.41 to 0.61). Nicotine-replacement therapy exposure was also associated with a decreased risk of intubation or death in hospitalized individuals with Covid-19 (13 vs. 73 patients, hazard ratio, 0.31; 95% CI, 0.17 to 0.57) but with an increased risk of all-cause mortality (251 vs. 231 deaths, hazard ratio, 1.49; 95% CI, 1.24 to 1.80).

In the second analysis, 128,768 individuals with major smoking-related diseases exposed to nicotine-replacement therapy were matched with 243,793 unexposed individuals. Individuals were aged on average 55.3 years (standard deviation: 11.4) and 53.3% were male. In the main multivariable analysis, nicotine-replacement therapy exposure was neither associated with risk of hospitalization with Covid-19 (240 patients in the nicotine-replacement therapy group and 398 patients in the unexposed group, hazard ratio, 1.13; 95% CI, 0.94 to 1.38) nor with risk of death or an intubation in hospitalized individuals with Covid-19 (48 vs. 61 patients, hazard ratio, 1.00; 95% CI, 0.65 to 1.54). All-cause mortality was higher in the nicotine-replacement therapy group (1040 vs. 366 deaths, hazard ratio, 3.83; 95% CI, 3.41 to 4.31).

**Conclusions:** This large-scale observational study suggests that smoking, measured by exposure to nicotine-replacement therapy, was associated with an increased risk of overall mortality during the first wave of SARS-CoV-2 epidemic in France, although it was associated with a lower risk of severe Covid-19 in individuals without major related-smoking diseases. Experimental and clinical studies are needed to disentangle the potential mechanisms of nicotine and/or smoking in Covid-19 risk. Whatever the nature of these associations, the global impact of smoking is harmful for health even over a short epidemic period.

## Introduction

Since the first cases were reported in December 2019 in Wuhan, China, infection with the severe acute respiratory coronavirus 2 (SARS-CoV-2) has become a worldwide pandemic. The virus has spread rapidly with more than 15 million people infected (and 620,000 deaths) as of July 23, 2020^1^. By the same date, 210 000 patients had tested positive for Covid-19 in France and more than 30 000 patients had died of Covid-19 (19 500 in-hospital deaths and 10 500 deaths in retirement homes and long-term care facilities)^2^.

The limitation of epidemic spread is currently based on nonpharmacologic interventions, including social distancing, the use of face masks, environmental hygiene, and hand washing^3^. To date, no pharmacologic intervention has shown any preventive effect on the spread of the epidemic.

Several Chinese studies have reported an unexpectedly low prevalence of current smoking among hospitalized patients with Covid-19^4^. Indeed, the rate of smokers in these studies ranged from 1.4 % to 12.6 %^4^. In this context, research has then focused on the relationship between smoking and Covid-19 and a recent systematic review, based on eighteen studies conducted in US, China, South Korea and Japan, confirmed the unusually low prevalence of current smoking among hospitalized Covid-19 patients. Smoking prevalence was less than one-fourth the expected prevalence based on gender-adjusted population smoking rates^5^. Considering those results, hypotheses of a potential protective effect of the nicotine or smoking on Covid-19 infection have emerged and several mechanisms were alleged^5–7^ However, these studies mostly compared observed to expected rates of smoking without direct comparison with individual controls. In addition, they included limited number of subjects and smoking information was mainly collected from medical records of hospitalized patients.

In this study, we used nicotine-replacement therapy, as a surrogate of smoking, in a nationwide, retrospective, matched “exposed/unexposed” cohort study using information from the French national health data system (SNDS, formerly SNIIRAM)^9,10^. The objective of this report is to examine the association of nicotine-replacement therapy as a surrogate of smoking with risk of hospitalization with Covid-19 and all-cause mortality in France.

## Methods

### Setting and data sources

The SNDS covers the entire French population, i.e. 67 million inhabitants. In the SNDS database, an anonymous unique individual identifier links, since 2006, information from two principal data sources: DCIR (the national health insurance claims database) and PMSI (the national hospital and discharge database)^9,10^.

The DCIR database includes individual information on outpatient medical care, laboratory tests, and reimbursed drugs. Drugs are coded according to the Anatomical Therapeutic Chemical (ATC) classification. The health expenditure of patients with long-term diseases (LTDs), such as cancer, diabetes, etc., is fully reimbursed, and their diagnosis is recorded according to the International Statistical Classification of Diseases and Related Health Problems, Tenth Revision (ICD-10). The DCIR database also collects patient data such as age, gender, vital status (and date of death when applicable) and eligibility for complementary universal health insurance (CMU-C), which provides free access to healthcare for low-income people^9,10^.

The PMSI database contains details of all admissions and accident and emergency attendances at all public and private hospitals in France. It contains dates of hospital admission and discharge, discharge diagnoses coded according to ICD-10 and type of medical or surgical acts coded using the French common classification of medical procedures (Classification Commune des Actes Médicaux [CCAM]).

The SNDS has been extensively used in France to conduct real-life studies, especially concerning the use, safety and efficacy of drugs. Many pharmaco-epidemiological studies have been based on the use of this database were conducted by our team EPI-PHARE^9–18^.

EPI-PHARE is a French Scientific Interest Group which has been created at the end of 2018 by two public institutions: the French National Agency for the Safety of Medicines and Health Products (ANSM) and the French National Health Insurance (CNAM).^19^ EPI-PHARE is the merging of both Department of Epidemiology of Health Products from ANSM and Department of Studies in Public Health from CNAM, which used to collaborate together since 10 years and have already published several articles in the field of pharmaco-epidemiology^9–18,20,21^. EPI-PHARE has been created to conduct pharmaco-epidemiological studies using complex and massive data from the SNDS to enlighten public authorities in their decision-making. EPI-PHARE has a regulatory permanent access to the data from the SNDS. This permanent access is given according the French Decree No. 2016-1871 of December 26, 2016 relating to the processing of personal data called “National Health Data System”^22^ and French law articles Art. R. 1461-13^23^ and 14^24^.

### Nicotine-replacement therapy and smoking data

In this study, we assume that almost all individuals with nicotine-replacement therapy exposure, at the entry study, were current smokers (for incident users) or recent quitters. Nicotine-replacement therapy (Code ATC N07BA01) has been reimbursed in France, when prescribed by a health professional, since January 1, 2019, whatever the number of prescriptions. Before this date, individuals who wanted to quit smoking were reimbursed, in 2107 and 2018, only one annual lump sum price of 150 euros for nicotine-replacement therapy (and 50 euros before 2017). The reimbursed forms include patch, gun, lozenge and tablets. No brand of oral spray or inhalator are reimbursed. Nasal spray is currently not available in France. More than 87 brand names are reimbursed in France (Supplementary Appendix, Table S1). Data on smoking status were absent from the database, but some information about medical care related to tobacco use was present (smoking use disorders): a hospital discharge diagnosis related to tobacco use since 2006 (ICD-10 codes F17, Z71.6 and Z72.0) or dispensing the annual lump sum price of nicotine-replacement therapy or dispensing varenicline (reimbursed since November 2017). Bupropion is not reimbursed in France.

### Exposed and unexposed groups to nicotine-replacement therapy

We conducted two separated analyses: the first one in individuals exposed to nicotine-replacement therapy without major smoking-related diseases (cancer, cardiovascular and/or respiratory diseases) and the second one in those presenting these conditions. Major smoking-related diseases were defined by the presence, in the 5 years prior to the study entry, of cancer (all types), cardiovascular diseases (myocardial infraction, heart failure or severe arrhythmia), stroke or respiratory diseases (individuals who had chronic obstructive pulmonary disease and/or asthma or those who had at least 3 reimbursements within the 12-month period preceding the study entry drugs for obstructive airway diseases (ATC R03). (Table S2 in the appendix).

In the first analysis, we included all individuals, aged between 18 and 75 years, who had been reimbursed at least one nicotine-replacement therapy between November 15, 2019, and February 15, 2020. We excluded individuals who had major smoking-related diseases.

For each exposed individual, we randomly selected, from the entire French Metropolitan population, up to two non-exposed individuals matched for the following variables: age (same year of birth), sex (male or female), department of residence (n=96 in France Metropolitan), and complementary universal health insurance (CMU-C). The same exclusion criteria were applied to non-exposed. In addition, individuals with smoking-use disorders were excluded from the non-exposed group. When a selected non-exposed individual had one or more exclusion criteria, a new one was randomly chosen and so on until the inclusion of individuals without criteria of exclusion.

In the second analysis, we included all individuals with major smoking-related diseases, aged between 18 and 75 years, who had been reimbursed at least one nicotine-replacement therapy between November 15, 2019, and February 15, 2020. For each exposed individual, we randomly selected, from the entire French Metropolitan population, up to two non-exposed individuals matched for the following variables: age, sex, department of residence, and CMU-C. Individuals with smoking-use disorders were excluded from the non-exposed group.

### Variables Assessed

Although we used complementary universal health insurance as a matching variable, we also used the social deprivation index in the analysis as another socioeconomic status variable^25^. This index measures the level of deprivation in individuals’ municipality of residency based on the area median household income, the percentage of high school graduates in the population aged 15 years and older, the percentage of manual workers in the active population, and unemployment. This index is divided into quintiles: the lower quintile (Q1) represents the least deprivation and the highest one (Q5) the most deprivation

Diabetes was defined by at least three reimbursements of antidiabetic drugs in the previous 12 months or long-term disease for diabetes (LTD for diabetes). As BMI is not recorded in the database, obesity was defined based on the use of antiobesity products or obesity-related hospitalization. Similarly, alcohol use disorders was identified based upon LTD or hospitalization diagnoses or reimbursements of specific drugs. History of substance abuse was based upon opiate substitution reimbursement or diagnosis related to substance abuse. Attempted suicide definition was based on hospital discharge in the 5 years prior to the study entry with ICD-10 X60-X84. For further details about variable definitions, see Table S3 in the Supplementary Appendix.

Consumption of antidepressants, benzodiazepines, hypnotics and neuroleptics drugs, anti-hypertensive drugs, lipid lowering drugs, and oral corticosteroids were each defined by the existence of at least 3 reimbursements within the previous 12 months (Table S3).

### Hospitalization with Covid-19

Information on hospital stays is usually reported in the PMSI with varying delays depending on the hospital establishment. Information on all hospital stays is integrated to SNDS monthly (with a few months later) and, for a given year, the information is definitively integrated to SNDS once a year (usually on July the following year). However, the French government encouraged, on April 2020, the hospital establishments to report in an exceptional and accelerated way (weekly or bi-monthly) information on the Covid-19 hospital stays (called PMSI “fast-track”)^26^. The report of information for hospitalization with Covid-19 was coordinated by a French governmental agency ATIH (Agence technique de l’information sur l’hospitalisation). Our study was based on data available on hospital discharges at June 7, 2020. At this date, 86,623 patients with hospitalization with Covid-19 were reported and identified, including 83,445 (96%) whose information could be linked with reimbursement information (i.e. without error on the anonymous identifier number). Of the 83,445 patients, 14,566 (17.5%) died in hospitals.

### End Points

The first end point was the time from study entry to hospitalization with Covid-19, defined as February 15, 2020. The second end point was the time from study entry to intubation or death during a hospital stay for Covid-19. As during the hospital stay, the date of death was available but not the date of intubation, we have chosen the date of hospitalization for intubated patients. For patients who died after intubation, the timing of the second end point was defined as the time of intubation. The third end point was the time from study entry to death whatever the cause (all-cause mortality). Follow-up for hospitalization with Covid-19, for intubation or death during a hospital stay for Covid-19 or for all-cause mortality continued through June 7, 2020.

### Statistical Analysis

Bivariate associations between treatment group and the measured patient characteristics were described and assessed with χ2 tests for categorical variables and the t-test for continuous ones. We also assessed the associations of the three end points (hospitalization with Covid-19, intubation or death during a hospital stay for Covid-19 and all-cause mortality) with patient characteristics. Conditional Cox proportional-hazards regression models were used to estimate the association between nicotine-replacement therapy exposure and each of the three end points. An initial multivariable conditional Cox regression model included matching factors and social deprivation index, obesity, diabetes, dementia, alcohol uses disorders, substance abuse, suicide attempt, anti-hypertensive drugs, lipid lowering drugs, antidepressants, benzodiazepines, hypnotics and neuroleptics drugs, nonsteroidal anti-inflammatory drugs and oral corticosteroids. In addition, to help account for the nonrandomized treatment of nicotine-replacement therapy, we used propensity-score methods to reduce the effects of confounding. The individual propensities for exposure to nicotine-replacement therapy were estimated with the use of a multivariable logistic-regression model that included the same covariates as the conditional Cox regression model. Associations between nicotine-replacement therapy exposure and the three end points were then estimated by multivariable Cox regression models with the use of two propensity score methods^27^.

The primary and main analysis used inverse probability weighting. In this analysis, the predicted probabilities from the propensity-score model were used to calculate the stabilized inverse-probability-weighting weight. To assess the balance of individual covariates before and after inverse probability of treatment weighting, standardized differences were calculated as the difference in means or proportions divided by the pooled standard deviation. The negligible difference was defined as an absolute standardized difference of less than 0.1^10,28^. We conducted a secondary analysis that included the inverse probability weighting and covariates. Conditional Fine-Gray competing risks analyses were also performed to account for the competing risk between all-cause mortality and hospitalization with Covid-19.

To understand the potential for an unmeasured confounder to render apparent significant ratio measures below 1.0 to be nonsignificant, when adjusted measures were found to be statistically significant we computed the E-value for the higher bound of the confidence interval^29,30^. All calculations were performed using SAS, version 9.4 software (SAS Institute Inc.).

## Results

During the study period, we identified 427,095 individuals with exposure to nicotine-replacement therapy, of whom 297,815 (69.7%) were without major smoking-related diseases and 129,280 (30.3%) with major smoking-related diseases (Figure 1).

**Figure 1.**
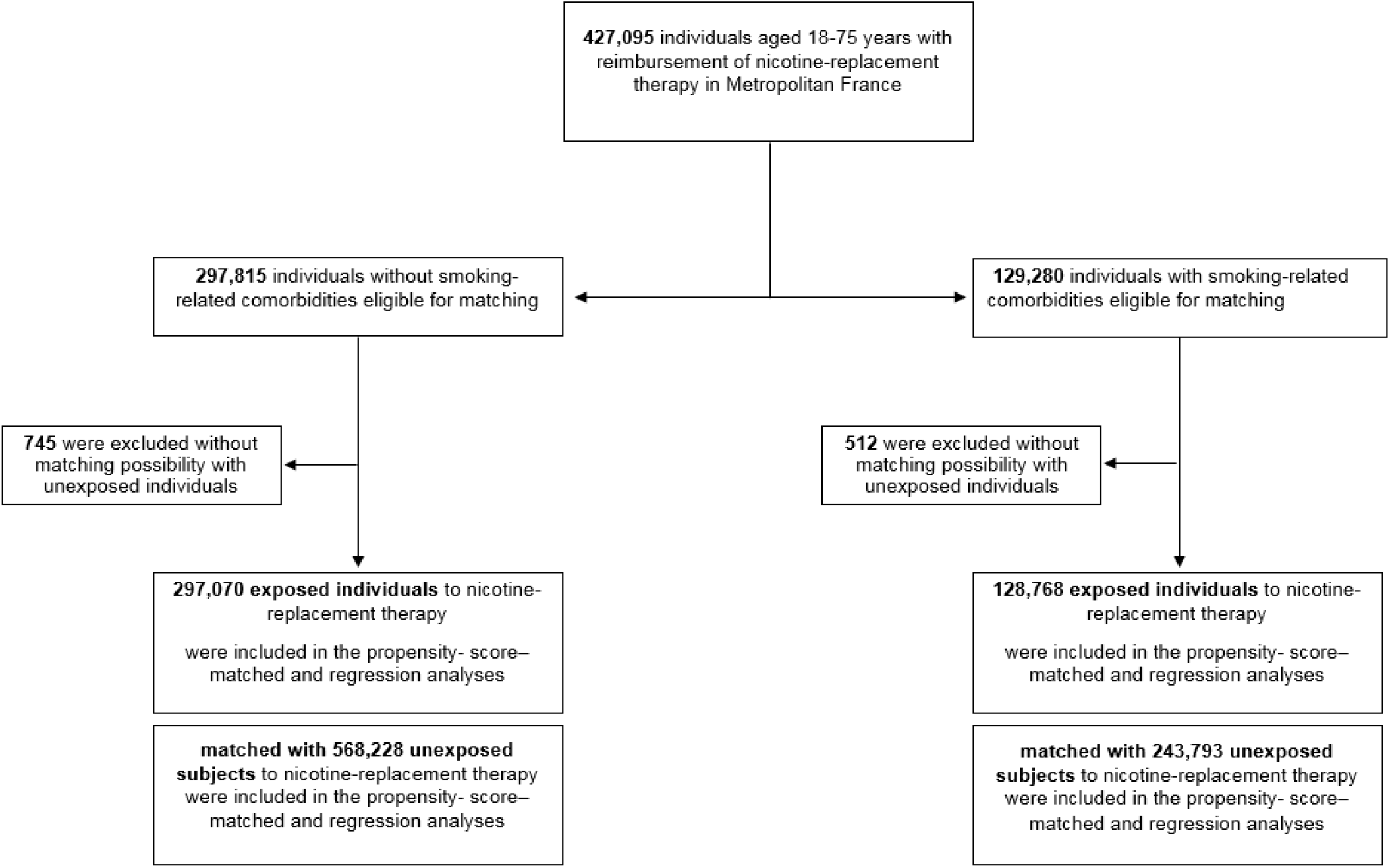
Study Cohort.

### Exposed and unexposed groups to nicotine-replacement therapy in individuals without major smoking-related diseases

Of 297,815 eligible exposed individuals to nicotine-replacement therapy, 297,070 could be matched (1:2) with 568,228 unexposed individuals (271,158 matched to two unexposed individuals, 25,912 with only one and 745 without matching).

The distribution of the patients’ baseline characteristics according to nicotine-replacement therapy exposure is shown in Table 1. Individuals were aged on average 45.6 years (standard deviation: 12.9) and 48.8% were male. Individuals exposed to nicotine-replacement therapy were more likely to be obese and to have diabetes, alcohol use disorders and substance abuse and more likely to have attempted suicide compared to unexposed individuals. All studied medications were more frequent, particularly psychiatric drugs, in the nicotine-replacement therapy (Table 1).

**Table 1.** Baseline characteristics of population with or without smoking-related diseases according to nicotine-replacement therapy exposure.

| Characteristic | Exposed individual without smoking-related diseases* |  |  | Exposed individual with smoking-related diseases** |  |  |
| --- | --- | --- | --- | --- | --- | --- |
|  | Exposed to nicotine replacement therapy<br>(N = 297,070) | Unexposed to nicotine-replacement therapy<sup>□</sup><br>(N = 568,228) | p-value | Exposed to nicotine replacement therapy<br>(N = 128,768) | Unexposed to nicotine replacement therapy<sup>□</sup><br>(N = 243,793) | p-value |
| **Sociodemographic** |  |  |  |  |  |  |
| Male sex – no. (%) | 144,955 (48.8) | 276,847 (48.7) | -<sup>□</sup> | 69,157 (53.7) | 129,722 (53.2) | -<sup>□</sup> |
| Age (years) – means ± SD | 45.6 ± 12.7 | 45.5 ± 12.7 | -<sup>□</sup> | 55.3 ± 11.3 | 55.3 ± 11.4 | -<sup>□</sup> |
| Age – no. (%) |  |  | -<sup>□</sup> |  |  | -<sup>□</sup> |
| 18 – 39 | 100,857 (34.0) | 193,656 (34.1) |  | 13,238 (10.3) | 25,221 (10.3) |  |
| 40 – 49 | 76,175 (25.6) | 146,107 (25.7) |  | 21,112 (16.4) | 40,037 (16.4) |  |
| 50 – 59 | 74,939 (25.2) | 143,076 (25.2) |  | 42,984 (33.4) | 80,867 (33.2) |  |
| 60 – 75 | 45,099 (15.2) | 85,389 (15.0) |  | 51,434 (39.9) | 97,668 (40.1) |  |
| Regions – no. (%)<sup>†</sup> |  |  | -<sup>□</sup> |  |  | -<sup>□</sup> |
| Ile de France<sup>§</sup> | 38,981 (13.1) | 75,974 (13.4) |  | 16,470 (12.8) | 31,729 (13.0) |  |
| Centre-Val de Loire | 11,749 (4.0) | 22,405 (3.9) |  | 4,991 (3.9) | 9,399 (3.9) |  |
| Bourgogne Franche Comté | 13,214 (4.4) | 25,178 (4.4) |  | 5,473 (4.3) | 10,355 (4.2) |  |
| Normandie | 18,031 (6.1) | 34,200 (6.0) |  | 7,748 (6.0) | 14,566 (6.0) |  |
| Hauts de France | 33,747 (11.4) | 63,595 (11.2) |  | 14,822 (11.5) | 27,676 (11.4) |  |
| Grand Est | 23,014 (7.7) | 43,688 (7.7) |  | 11,441 (8.9) | 21,466 (8.8) |  |
| Pays de Loire | 16,496 (5.6) | 31,628 (5.6) |  | 6,248 (4.9) | 11,912 (4.9) |  |
| Bretagne | 18,054 (6.1) | 34,567 (6.1) |  | 7,178 (5.6) | 13,609 (5.6) |  |
| Nouvelle Aquitaine | 31,738 (10.7) | 60,519 (10.7) |  | 13,892 (10.8) | 26,196 (10.7) |  |
| Occitanie | 32,469 (10.9) | 62,219 (10.9) |  | 14,624 (11.4) | 27,764 (11.4) |  |
| Auvergne-Rhône Alpes | 35,542 (12.0) | 67,960 (12.0) |  | 14,847 (11.5) | 28,083 (11.5) |  |
| Provence Alpes Cote d'Azur | 22,678 (7.6) | 43,668 (7.7) |  | 10,406 (8.1) | 19,829 (8.1) |  |
| Corse | 1,357 (0.5) | 2,627 (0.5) |  | 628 (0.5) | 1,209 (0.5) |  |
| CMU_C – no. (%) | 39,315 (13.2) | 73,177 (12.9) | -<sup>□</sup> | 19,194 (14.9) | 35,119 (14.4) | -<sup>□</sup> |
| Social deprivation index (quintiles) |  |  | <0.0001 |  |  | <0.0001 |
| 1 (the least deprivation) | 49,203 (16.6) | 101,849 (17.9) |  | 19,215 (14.9) | 41,992 (17.2) |  |
| 2 | 57,363 (19.3) | 112,922 (19.9) |  | 23,065 (17.9) | 46,767 (19.2) |  |
| 3 | 61,603 (20.7) | 115,548 (20.3) |  | 26,636 (20.7) | 49,337 (20.2) |  |
| 4 | 62,421 (21.0) | 114,363 (20.1) |  | 27,813 (21.6) | 49,909 (20.5) |  |
| 5 (the most deprivation) | 62,050 (20.9) | 113,229 (19.9) |  | 30,378 (23.6) | 51,551 (21.1) |  |
| Unknown | 4,430 (1.5) | 10,317 (1.8) |  | 1,661 (1.3) | 4,237 (1.7) |  |
| **Comorbidities – no. (%)** |  |  |  |  |  |  |
| Cardiovascular disease<sup>a</sup> | -* | -* |  | 41,185 (32.0) | 16,514 (6.8) |  |
| Respiratory disease<sup>b</sup> | -* | -* |  | 77,976 (60.6) | 16,518 (6.8) |  |
| Cancer | -* | -* |  | 34,670 (26.9) | 17,942 (7.4) |  |
| Obesity | 8,892 (3.0) | 5,979 (1.1) | <0.0001 | 22,884 (17.8) | 21,409 (8.8) | <0.0001 |
| Diabetes | 14,423 (4.9) | 22,467 (4.0) | <0.0001 | 17,346 (13.5) | 21,074 (8.6) | <0.0001 |
| Dementia | 167 (0.1) | 314 (0.1) | 0.86 | 299 (0.2) | 623 (0.3) | 0.17 |
| Alcohol use disorders | 23,555 (7.9) | 8,113 (1.4) | <0.0001 | 21,434 (16.6) | 4,818 (2.0) | <0.0001 |
| Substance abuse | 11,504 (3.9) | 4,109 (0.7) | <0.0001 | 7,018 (5.5) | 1,531 (0.6) | <0.0001 |
| Suicide attempt | 6,529 (2.2) | 2,766 (0.5) | <0.0001 | 3,993 (3.1) | 1,112 (0.5) | <0.0001 |
| **Medications – no. (%)** |  |  |  |  |  |  |
| Anti-hypertensive drugs<sup>c</sup> |  |  | <0.0001 |  |  | <0.0001 |
| ACE inhibitors | 12,319 (4.1) | 17,667 (3.1) |  | 23,026 (17.9) | 17,826 (7.3) |  |
| Other anti-hypertensive drugs | 31,722 (10.7) | 52,312 (9.2) |  | 35,735 (27.8) | 46,757 (19.2) |  |
| Lipid lowering drugs<sup>c</sup> | 27,589 (9.3) | 31,278 (5.5) | <0.0001 | 43,894 (34.1) | 34,992 (14.4) | <0.0001 |
| Psychiatric drugs<sup>c</sup> | 127,780 (43.0) | 110,077 (19.4) | <0.0001 | 75,443 (58.6) | 59,044 (24.2) | <0.0001 |
| Antidepressants<sup>c</sup> | 48,812 (16.4) | 35,258 (6.2) | <0.0001 | 32,808 (25.5) | 20,316 (8.3) | <0.0001 |
| Anxiolytics<sup>c</sup> | 52,343 (17.6) | 31,556 (5.6) | <0.0001 | 39,647 (30.8) | 20,126 (8.3) | <0.0001 |
| Hypnotics<sup>c</sup> | 18,380 (6.2) | 9,403 (1.7) | <0.0001 | 16,661 (12.9) | 7,113 (2.9) | <0.0001 |
| Neuroleptics<sup>c</sup> | 21,475 (7.2) | 9,315 (1.6) | <0.0001 | 11,913 (9.3) | 4,725 (1.9) | <0.0001 |
| NSAID<sup>c</sup> | 36,777 (12.4) | 44,532 (7.8) | <0.0001 | 16,002 (12.4) | 22,196 (9.1) | <0.0001 |
| Oral corticosteroids<sup>c</sup> | 7,631 (2.6) | 8,141 (1.4) | <0.0001 | 13,314 (10.3) | 6,239 (2.6) | <0.0001 |

ACE Angiotensin-converting enzyme; CMU\_C complementary universal health insurance; NRT Nicotine replacement therapy; NSAID Nonsteroidal anti-inflammatory drug; SD Standard deviation
\*Cardiovascular history, respiratory history and cancer history were considered as smoking-related comorbidities. Exposed and unexposed to nicotine replacement therapy individual have no smoking-related comorbidities.
\*\*All exposed individual to nicotine replacement therapy person have smoking related comorbidities whereas unexposed individual can have them or not.
‡Unexposed population was matched by age, sex, departments and CMU\_C.
†Results are displayed by regions but unexposed population was matched by departments (more precise).
§Paris and great Paris.
<sup>a</sup>Heart failure, ischemic heart disease, arrhythmia and stroke.
<sup>b</sup>Chronic obstructive pulmonary disease, asthma or bronchodilators use.
<sup>c</sup>At least three dispensations in previous year.

In the nicotine-replacement therapy group, 140,795 individuals (47.4%) were incident users. The average number of reimbursements in the previous six months was 2.6 (standard deviation: 2.6). 19.9% had two reimbursements and 32.7% had three reimbursements and more (Table 2). For the last reimbursement, the form was a patch for 75.5% of individuals, a lozenge for 22.9% and a gum for 22.1%. 27.2% had simultaneously a patch and one other non-patch form (Table 2).

**Table 2.** Description of nicotine-replacement therapy exposure on population with or without smoking-related diseases.

|  | Exposed individual without<br>smoking-related diseases*<br>N= 297,070 | Exposed individual with<br>smoking-related diseases**<br>N= 128,768 |
| --- | --- | --- |
| Type of users – *n* (%) |  |  |
| Incident | 140,795 (47.4) | 44903 (34.9) |
| Prevalent<sup>a</sup> | 156,275 (52.6) | 83865 (65.1) |
| Number of reimbursements<sup>§</sup> – mean<br>± SD | 2.6 ± 2.6 | 3.2 ± 2.8 |
| Number of reimbursements<sup>§</sup> – *n* (%) |  |  |
| 1 | 140,795 (47.4) | 44903 (34.9) |
| 2 | 59,036 (19.9) | 24775 (19.2) |
| ≥3 | 97,239 (32.7) | 59090 (45.9) |
| Date of last reimbursement – *n* (%) |  |  |
| February 2020 | 80,531 (27.1) | 37777 (29.3) |
| January 2020 | 113,896 (38.3) | 50569 (39.3) |
| December 2019 | 65,959 (22.2) | 26984 (21.0) |
| November 2019 | 36,684 (12.4) | 13438 (10.4) |
| Type of form – *n* (%)<sup>†b</sup> |  |  |
| Patch | 224,192 (75.5) | 96184 (74.7) |
| Gum | 65,506 (22.1) | 23992 (18.6) |
| Lozenge | 67,982 (22.9) | 30562 (23.7) |
| Tablets | 21,447 (7.2) | 9232 (7.2) |
| Type of form – *n* (%)<sup>b</sup> |  |  |
| Patch only | 144,655 (48.7) | 65970 (51.2) |
| Patch + Non-patch form | 80,933 (27.2) | 30214 (23.5) |
| Non-patch form only | 71,482 (24.1) | 32584 (25.3) |

\*Cardiovascular history, respiratory history and cancer history were considered as smoking-related comorbidities. Exposed and unexposed to nicotine replacement therapy individual have no smoking-related comorbidities.
\*\*All exposed individual to nicotine replacement therapy person have smoking related comorbidities whereas unexposed individual can have them or not.
<sup>a</sup>No nicotine replacement therapy reimbursement in the 6 months preceding the last reimbursement.
<sup>b</sup>Last nicotine replacement therapy reimbursement.
<sup>§</sup>In the previous six months.
<sup>†</sup>A person can take several types of forms: sum of percentage may exceed 100%.

The odds ratios (with 95% confidence intervals) for the association between exposure to nicotine-replacement therapy and all the variables included in the propensity-score model are shown in Table S4. After inverse probability of treatment weighting, baseline characteristics of individuals according to nicotine-replacement therapy exposure were well balanced (Tables S5-6).

From February 15, 2020 to June 7, 2020, the first end point (hospitalization with Covid-19) occurred in 647 patients (151 patients in the nicotine-replacement therapy group and 496 patients in the unexposed group). Kaplan-Meier survival curves of time to hospitalization for Covid-19 stratified by exposure group are shown in Figure S1. In the crude, unadjusted analysis, individuals exposed to nicotine-replacement therapy were less likely to have had a first endpoint event than those who were not exposed (hazard ratio, 0.58; 95% CI, 0.49 to 0.70) (Table 3). In the main multivariable analysis with inverse probability weighting according to the propensity score, nicotine-replacement therapy exposure was again associated with a decreased risk of hospitalization with covid-19 (hazard ratio, 0.50; 95% CI, 0.41 to 0.61). The E-value of the lower bound of confidence interval was 2.64. Additional multivariable propensity-score analysis yielded similar results (Table 3). Results from Fine-Gray models were similar to those with the cause-specific conditional Cox proportional hazards regression model (hazard ratio, 0.49; 95% CI 0.41 – 0.59). The negative association of nicotine-replacement therapy with hospitalization with Covid-19 was observed in all subgroups according to sex, age and patterns of nicotine replacement therapy exposure (Table 4). This association tended, however, to be more pronounced in male (hazard ratio, 0.42; 95% CI, 0.32 to 0.55), in individuals aged 45 years or more (hazard ratio, 0.47; 95% CI, 0.37 to 0.59), in incident users (hazard ratio, 0.43; 95% CI, 0.32 to 0.58) and in individuals with simultaneous reimbursement of patch and one another non-patch form (hazard ratio, 0.40; 95% CI, 0.26 to 0.61). The association was less pronounced in individuals with more than three reimbursements (0.77; 95% CI, 0.54 to 1.09).

**Table 3.** Associations between nicotine-replacement therapy and hospitalization with Covid-19, death with COVID-19 or intubation and all-cause mortality in the Crude Analysis, Multivariable Analysis, and Propensity-Score Analyses.

| Analysis | Hospitalization with Covid-19 | Death with COVID-19 or intubation | All-cause mortality |
| --- | --- | --- | --- |
| **Exposed individual without smoking-related diseases<sup>a</sup>** |  |  |  |
| No. of events |  |  |  |
| Nicotine Replacement Therapy, n= 297,070 | 151 | 13 | 251 |
| No Nicotine Replacement Therapy, n= 568,228 | 496 | 73 | 235 |
| Crude analysis — hazard ratio (95% CI) | 0.58 (0.49 – 0.70) | 0.34 (0.19 – 0.62) | 2.01 (1.68 – 2.41) |
| Multivariable analysis — hazard ratio (95% CI)* | 0.49 (0.40 – 0.60) | 0.23 (0.11 – 0.49) | 1.38 (1.11 – 1.72) |
| Propensity-score analyses — hazard ratio (95% CI) |  |  |  |
| With inverse probability weighting † | 0.50 (0.41 – 0.61) | 0.31 (0.17 – 0.57) | 1.49 (1.24 – 1.80) |
| With inverse probability weighting and covariates‡ | 0.49 (0.40 – 0.60) | 0.25 (0.12 – 0.52) | 1.32 (1.07 – 1.63) |
| **Exposed individual with smoking-related diseases<sup>b</sup>** |  |  |  |
| No. of events |  |  |  |
| Nicotine Replacement Therapy, n= 128,768 | 240 | 48 | 1,040 |
| No Nicotine Replacement Therapy, n= 243,793 | 298 | 61 | 366 |
| Crude analysis — hazard ratio (95% CI) | 1.50 (1.26 – 1.78) | 1.48 (1.01 – 2.17) | 5.38 (4.78 – 6.07) |
| Multivariable analysis — hazard ratio (95% CI)* | 0.98 (0.78 – 1.24) | 0.69 (0.37 – 1.27) | 3.52 (3.01 – 4.12) |
| Propensity-score analyses — hazard ratio (95% CI) |  |  |  |
| With inverse probability weighting† | 1.13 (0.94 – 1.38) | 1.00 (0.65 – 1.54) | 3.83 (3.41 – 4.31) |
| With inverse probability weighting and covariates‡ | 0.93 (0.75 – 1.16) | 0.71 (0.40 – 1.25) | 3.39 (2.95 – 3.90) |

<sup>a</sup> Cardiovascular history, respiratory history and cancer history were considered as smoking-related comorbidities. Exposed and unexposed to nicotine-replacement therapy individual have no smoking-related comorbidities.
<sup>b</sup> All exposed individual to nicotine-replacement therapy person have smoking related comorbidities whereas unexposed individual can have them or not.
\*Shown is the hazard ratio from the multivariable Cox proportional-hazards model, with stratification according to sex, age, department and CMU-c, and with additional adjustment for social deprivation index, obesity, diabetes, dementia, alcohol uses disorders, substance abuse, suicide attempt, anti-hypertensive drugs, lipid lowering drugs, antidepressants, benzodiazepines, hypnotics and neuroleptics drugs, nonsteroidal anti-inflammatory drugs and oral corticosteroids.
† Shown is the primary analysis with a hazard ratio from the univariate Cox proportional-hazards model with inverse probability weighting according to the propensity score.
‡ Shown is the hazard ratio from the multivariable Cox proportional-hazards model with the same strata and covariates with inverse probability weighting according to the propensity score.

**Table 4.**
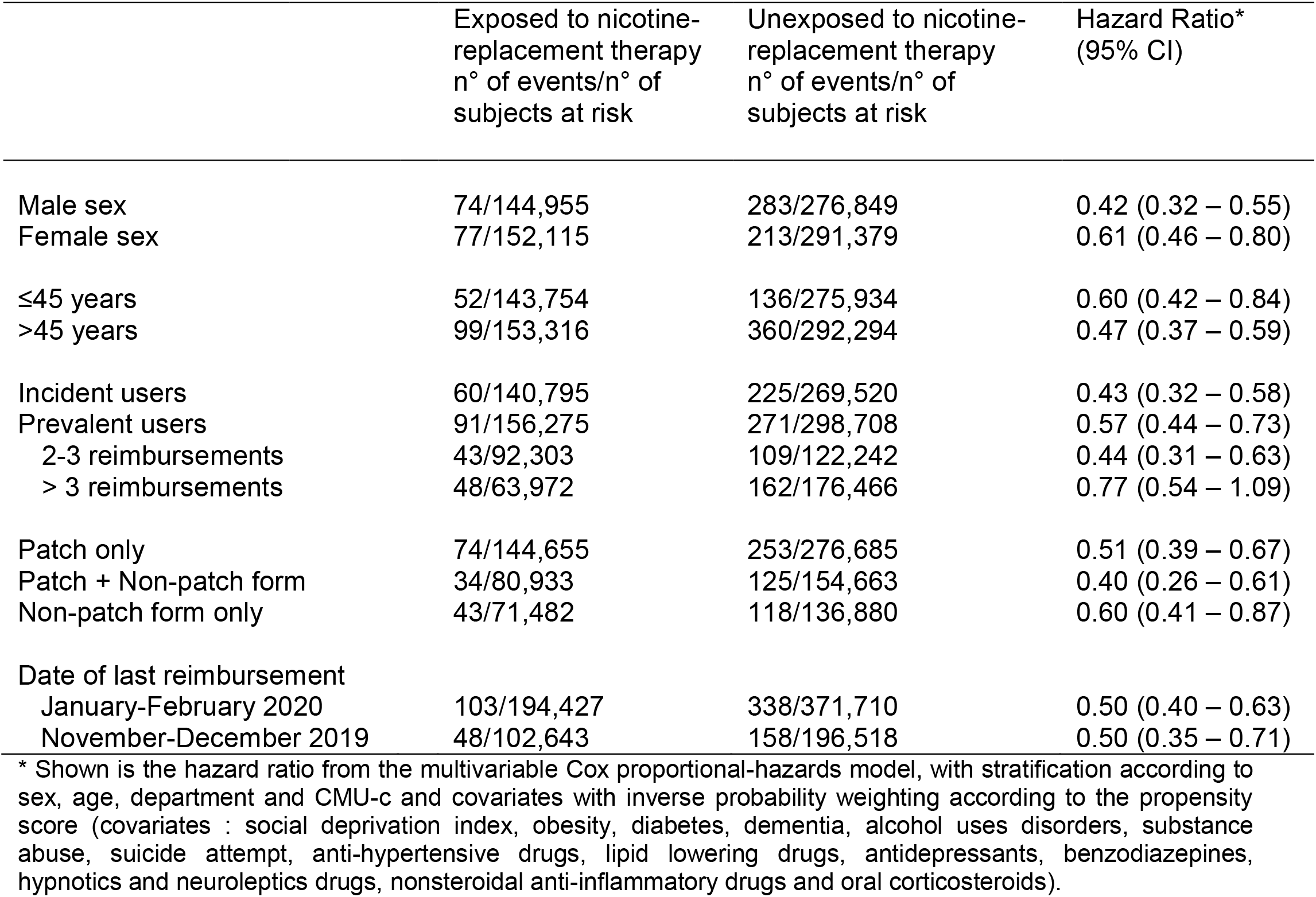
Associations between nicotine-replacement therapy and hospitalization with Covid-19 according to subgroups on population without smoking-related diseases.

|  | Exposed to nicotine-replacement therapy<br>n° of events/n° of subjects at risk | Unexposed to nicotine-replacement therapy<br>n° of events/n° of subjects at risk | Hazard Ratio*<br>(95% CI) |
| --- | --- | --- | --- |
| Male sex | 74/144,955 | 283/276,849 | 0.42 (0.32 – 0.55) |
| Female sex | 77/152,115 | 213/291,379 | 0.61 (0.46 – 0.80) |
| ≤45 years | 52/143,754 | 136/275,934 | 0.60 (0.42 – 0.84) |
| >45 years | 99/153,316 | 360/292,294 | 0.47 (0.37 – 0.59) |
| Incident users | 60/140,795 | 225/269,520 | 0.43 (0.32 – 0.58) |
| Prevalent users | 91/156,275 | 271/298,708 | 0.57 (0.44 – 0.73) |
| 2-3 reimbursements | 43/92,303 | 109/122,242 | 0.44 (0.31 – 0.63) |
| > 3 reimbursements | 48/63,972 | 162/176,466 | 0.77 (0.54 – 1.09) |
| Patch only | 74/144,655 | 253/276,685 | 0.51 (0.39 – 0.67) |
| Patch + Non-patch form | 34/80,933 | 125/154,663 | 0.40 (0.26 – 0.61) |
| Non-patch form only | 43/71,482 | 118/136,880 | 0.60 (0.41 – 0.87) |
| Date of last reimbursement |  |  |  |
| January-February 2020 | 103/194,427 | 338/371,710 | 0.50 (0.40 – 0.63) |
| November-December 2019 | 48/102,643 | 158/196,518 | 0.50 (0.35 – 0.71) |

\* Shown is the hazard ratio from the multivariable Cox proportional-hazards model, with stratification according to sex, age, department and CMU-c and covariates with inverse probability weighting according to the propensity score (covariates : social deprivation index, obesity, diabetes, dementia, alcohol uses disorders, substance abuse, suicide attempt, anti-hypertensive drugs, lipid lowering drugs, antidepressants, benzodiazepines, hypnotics and neuroleptics drugs, nonsteroidal anti-inflammatory drugs and oral corticosteroids).

In the main multivariable analysis with inverse probability weighting according to the propensity score, nicotine-replacement therapy use was both associated with a decreased risk of intubation or death in hospitalized individuals with Covid-19 (13 vs. 73 patients, hazard ratio, 0.31; 95% CI, 0.17 to 0.57) and with an increased risk of all-cause mortality (251 vs. 231 deaths, hazard ratio, 1.49; 95% CI, 1.24 to 1.80) (Table 3).

### Exposed group to nicotine-replacement therapy in individuals with major smoking-related diseases and unexposed group

Of 133,021 eligible exposed individuals to nicotine-replacement therapy, 128,768 could be matched (1:2) with 243,793 unexposed individuals (115,025 matched to two unexposed individuals, 13,743 with only one and 512 without matching). In the exposed group, 26.9% had cancer, 32.0% had cardiovascular/stroke diseases and/or 60.6% had respiratory diseases.

The distribution of the patients’ baseline characteristics according to nicotine-replacement therapy exposure is shown in Table 1. Individuals were aged on average 55.3 years (standard deviation: 11.4) and 53.3% were male.

In the nicotine-replacement therapy group, 44,903 individuals (34.9%) were incident users. The average number of reimbursements in the previous six months was 3.2 (standard deviation: 2.8) (Table 2).

Nicotine-replacement therapy use was associated with increased risks of the three end points in the crude analysis. However, in the main multivariable analysis, nicotine-replacement therapy group was neither associated with risk of hospitalization with Covid-19 (240 patients in the nicotine-replacement therapy group and 298 patients in the unexposed group, hazard ratio, 1.13; 95% CI, 0.94 to 1.38) nor with risk of death or an intubation in hospitalized individuals with Covid-19 (48 vs. 61 patients, hazard ratio, 1.00; 95% CI, 0.65 to 1.54). All-cause mortality was higher in the nicotine-replacement therapy group (1040 vs. 366 deaths, hazard ratio, 3.83; 95% CI, 3.41 to 4.31).

## Discussion

In this study based on a large nationwide, retrospective, matched “exposed/unexposed” cohort study followed during the period of the Covid-19 epidemics peak in France, exposure to nicotine-replacement therapy - a surrogate of current or recent smoking - was associated with a decreased risk of severe Covid-19, as measured by hospitalization with Covid-19 in individuals free of major smoking-related diseases (hazard ratio, 0.50; 95% CI, 0.41 to 0.61). Findings were consistently robust whatever the use of classical or more suitable models. Conversely, all-cause mortality was significantly higher in the nicotine-replacement therapy group (hazard ratio, 1.49; 95% CI, 1.24 to 1.80). In individuals with major smoking-related diseases, neither the risk of hospitalization with Covid-19 nor the risk in-hospital death or intubation in hospitalized patients with Covid-19 was associated with nicotine-replacement therapy. However, overall mortality in those with major smoking-related diseases exposed to nicotine-replacement therapy was markedly higher than in unexposed individuals (hazard ratio, 3.83; 95% CI, 3.41 to 4.31).

Several studies have reported a lower frequency of smoking among Covid-19 hospitalized patients compared to the general population^4,5,31^. Our findings provide further evidence of a negative association between smoking and severe Covid-19, though they suggest that such an association may be limited to individuals without major related-smoking conditions. Despite its strength, the nature (causal or not) of this association is not known and the observational design of this study prevents to drawing any conclusion about it. Even though this association would have been causal, it is not clear whether nicotine, other smoking components or smoking *per se* might be involved. It was hypothesized that nicotine might have a protective effect. One hypothesis concerns the implication of the nicotinic cholinergic system in the inflammatory syndrome of Covid-19^8^. Nicotine, a cholinergic agonist, has anti-inflammatory proprieties through the anti-inflammatory cholinergic pathway via 7α nicotinic acetylcholine receptors (7α nAChRs), inhibiting the expression of several pro-inflammatory cytokines such as IL-6, Il-1 or TNF^5^. In murine model, nicotine reduced lung inflammation acute respiratory distress syndrome^32^. Severe progression of Covid-19 infection has been associated with increased level of those cytokines, leading to cytokine storm with coagulation disturbance and multiorgan failure^33,34^. Finally, nicotine might upregulate angiotensin converting enzyme 2 (ACE2) expression, an enzyme used as a receptor by SARS-CoV-2 for cell entry^7^, but it is unknown if this effect is beneficial or detrimental for the patient.

In observational studies, assessing the effects of nicotine-replacement therapy on Covid-19 (or other conditions) independently of smoking is not obvious and the choice of the comparator group is particularly challenging. Choosing smokers, as a comparator group, is not entirely satisfactory as smokers are also exposed to nicotine. Choosing non-smokers or ex-smokers, as a comparator group, does not allow to disentangle the role of nicotine from other cigarette smoke compound. Similarly, the relationship of smoking with Covid-19, independently of nicotine, is also particularly difficult to investigate in observational studies as nicotine is a basic component of smoking.

Understanding the respective roles of nicotine and smoking, independently of nicotine, in their relationship with the Covid-19 is essential because the implications of the present findings may be opposite. Even if smoking independently of nicotine is implied, smoking-related risks in terms of morbidity and mortality consistently remain higher than potential benefits, even in individuals without smoking-related conditions despite their possibly decreased risk of severe Covid-19 suggested by our results (Table 3).

Cigarette smoking causes about one of every five deaths in the United States (480,000)^35^ and one of every six deaths in France (78,000) each year^36^. The major causes of excess mortality among smokers are diseases related to smoking, including cancers and respiratory and vascular diseases35,37,38

On the other hand, if nicotine proves to be pharmacologically implied in the negative association observed in individuals free of major smoking-related diseases, this could lead to consider nicotine-replacement therapy use as an option for smokers since it could/might also be useful for quitting smoking. However, the fact that the negative association was more pronounced in incident users of nicotine-replacement therapy group (mostly current smokers) than in prevalent users and more pronounced in those with three or more reimbursements (mostly ex-smokers or recent quitters) argues in favor of a role of nicotine rather than of smoking, unless the bronchial concentrations obtained by nicotine-replacement therapy are insufficient to reproduce the endobronchial effect of nicotine provided by tobacco smoke^39,40^. In vitro and in vivo experimental and clinical studies are necessary to better understand the potential physiopathological and pharmacological mechanisms of nicotine and/or smoking in Covid-19.

Our results assessed only the association of nicotine-replacement therapy with Covid-19 forms that required hospitalization. The results cannot be thus generalized to asymptomatic forms or to mild-to-moderate forms of Covid-19.

We used nicotine-replacement therapy as a surrogate of current or recent smoking. This marker has an excellent sensitivity and probably a very low specificity because the majority of current smokers do not use nicotine-replacement therapy. In the unexposed groups, the frequency of smokers was not known with precision. In France, daily smoking was reported by 25% of the general population in 2018 (28% of men and 23% of women)^41^. This rate is likely to be a bit lower among unexposed individuals without smoking use disorders (4%). This classification bias is likely to reduce the observed associations toward the null rather than artificially create associations.

Nicotine-replacement therapy can also be dispensed in France without reimbursement (over-the-counter). Only reimbursement data were available in our study. However, the rate of reimbursed products, from November 2019 to February 2020, is about 80% for the patch form and 50% for the other forms^42^. All reimbursed products in France were included in the study in the exposed groups. The use of nicotine-replacement therapy without reimbursement in the unexposed groups should be low because the two unexposed groups presented a small sample of the French population (less than 4% of adult French population) and the lack of data of exposure without reimbursement would not thus substantially modify the main results.

Smoking is associated with several illnesses including cancer, cardiovascular diseases and respiratory diseases which may modify the risk of hospitalization and prognosis of patients with Covid-19^43–45^. To study the specific role of smoking on Covid-19 risk under more controlled design, we conducted the first analysis in individuals without these conditions in both exposed and unexposed groups. On the other hand, to study the global smoking impact, we conducted an additional analysis of overall mortality in exposed individuals with and without related-smoking diseases.

It could be argued that smokers may have more limited access to hospitals and care facilities during the epidemic period due to socioeconomic factors. To take into account the potential confounding, exposed and unexposed individuals to nicotine-replacement therapy were matched by complementary universal health insurance which provides free access to healthcare for low-income people and an additional social indicator (social deprivation index) had been taken into account in all multivariable analyses. Only morbid obesity was assessed in this study and we did not have information on the body mass index values. However, diabetics without cardiovascular diseases (as in the population of our first analysis) are more frequently overweight and obese. Morbid obesity and diabetes were taken into account in all approaches of multivariate analyzes.

Residual confounding of effects of unmeasured or unknown factors could not be excluded. However, the E-value for the higher bound of the confidence interval was relatively high (2.64) which suggests that important unmeasured confounding would be needed to explain away the effect estimate of nicotine-replacement therapy on the risk of hospitalization with Covid-19 in individuals free of major smoking-related diseases.

For hospitalization with Covid-19, we used information reported from an exceptional and accelerated way coordinated by public French authorities^26^. Santé Publique France (the French center comparable to US Centers for Disease Control and Prevention) estimated that there were in France, between March 1, 2020 and June 9, 2020, 102,863 patients who were hospitalized with covid-19, including 18,912 patients who died in hospitals^46^. Of the 102,863 patients, 11,961 were still hospitalized on the date of June 9, 2020, including 955 patient in intensive care (with or without intubation)^2^. Before the date of March 1, 2020, there was less than 400 hospitalizations with Covid-19 in France. Therefore, our study included approximately 80% of all patients (83,455/102,863) who were hospitalized in France with COVID-19 and more than 90% if we exclude patients who are still hospitalized. The information was reported from 614 establishments from all French regions except two overseas regions, Martinique and Mayotte. 81% of patients were hospitalized in University hospitals and public general hospitals. Some hospital establishments did not transmit the files of patients with Covid-19, either by absence of cases, or by lack of technical support or time. Although we could not include all of the hospitalized patients in France, we can assume that the availability of hospitalization data was not related to use of nicotine-replacement therapy. We cannot exclude the fact that patients who were still hospitalized on June 7 might be related to nicotine-replacement therapy or to the characteristics of patients using it. However, the frequency of patients who were still hospitalized was relatively low (10%) and the analyses carried out on hospitalized patients in March 2020 gave similar results.

In conclusion, this large-scale observational study suggests that current or recent smoking, measured by exposure to nicotine-replacement therapy, was associated with an increased risk of overall mortality during the first wave of SARS-CoV-2 epidemic in France, although it was associated with a decreased risk of severe covid-19 in individuals without major related-smoking diseases. In vitro and in vivo experimental and clinical studies are needed to better understand the potential mechanisms of nicotine and/or smoking on Covid-19 risk. Whatever the nature of the associations between smoking and Covid-19 risk, the global impact of smoking is harmful for health, notably for overall mortality, even during a short epidemic period.

## Data Availability

According to data protection and the French regulation, the authors can not publicly release the data from the SNDS. However, any person or structure, public or private, for-profit or non-profit, is able to access SNDS data upon authorization from the CNIL, in order to carry out a study, a research or an evaluation of public interest (https://www.snds.gouv.fr)

## Acknowledgment

We thank Pr Joêl Ankri, Pr Bertrand Dautzenberg, Pr Loic Josseran, Pr Olivier Saint-Lary and Dr Jean-Marie Vailloud for very valuable advices.

## Supplementary Information

### Glossary

#### Table

**Table S1.**
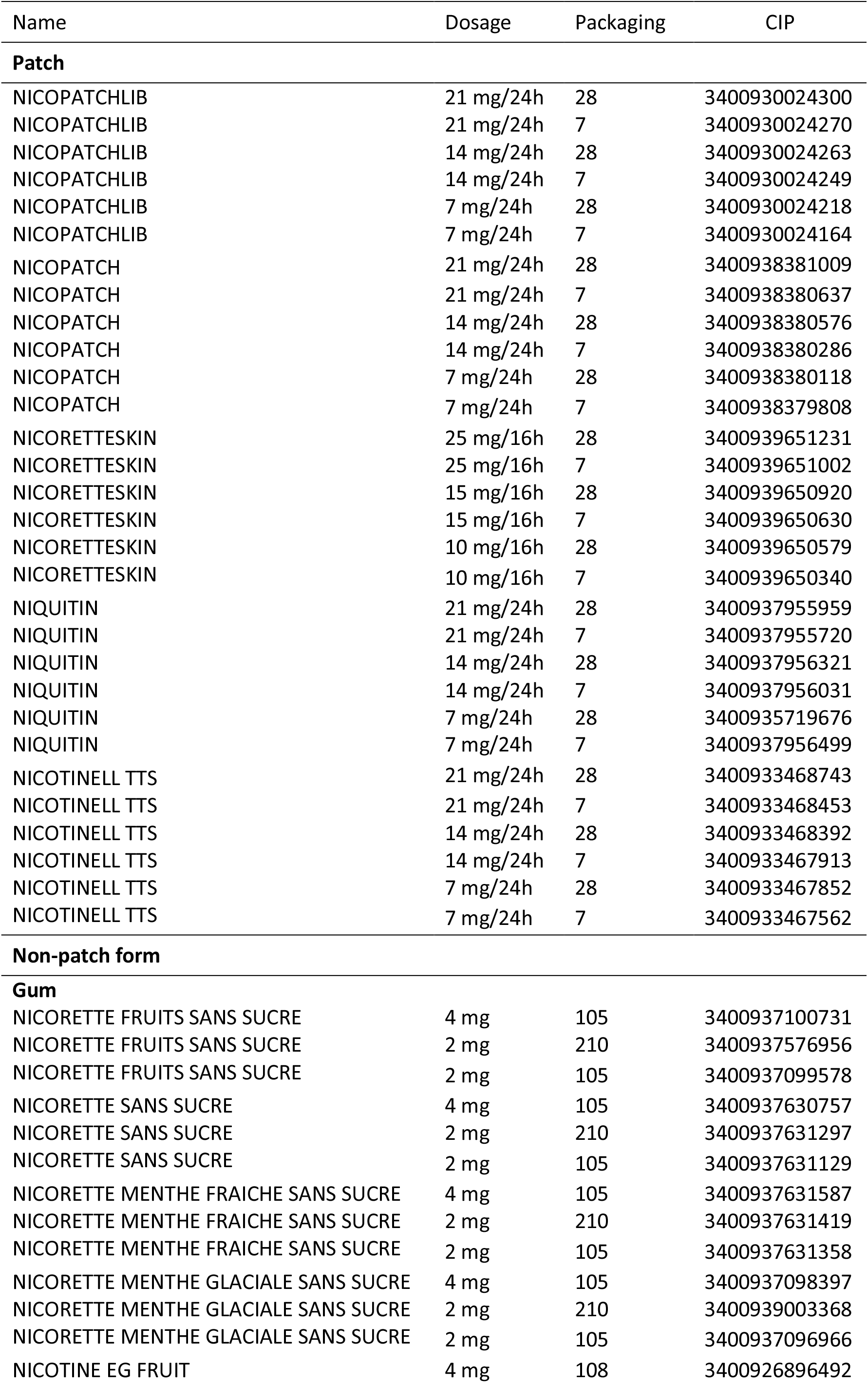
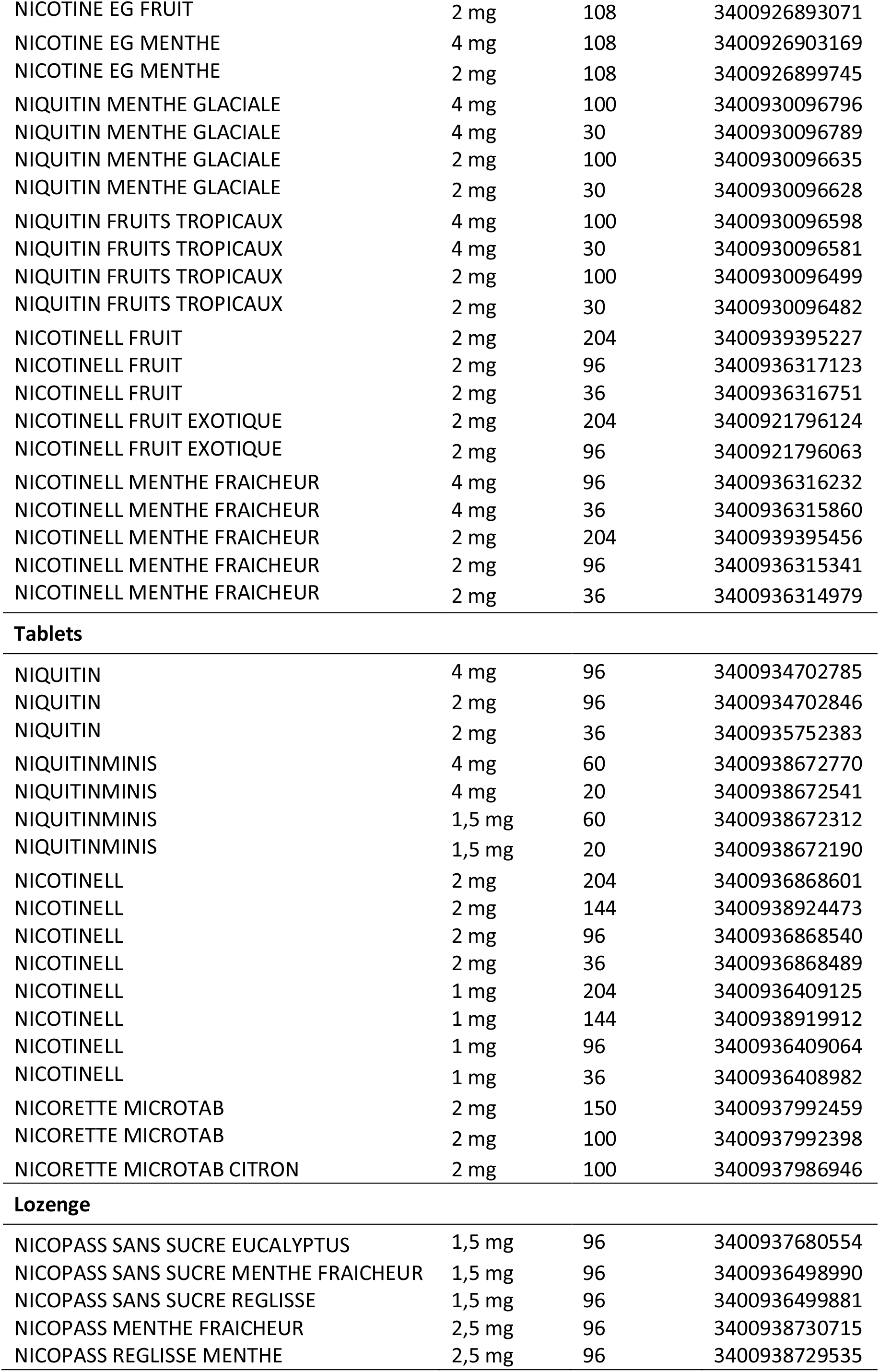
List of reimbursed nicotine-replacement therapy products in France.

| Name | Dosage | Packaging | CIP |
| --- | --- | --- | --- |
| **Patch** |  |  |  |
| NICOPATCHLIB | 21 mg/24h | 28 | 3400930024300 |
| NICOPATCHLIB | 21 mg/24h | 7 | 3400930024270 |
| NICOPATCHLIB | 14 mg/24h | 28 | 3400930024263 |
| NICOPATCHLIB | 14 mg/24h | 7 | 3400930024249 |
| NICOPATCHLIB | 7 mg/24h | 28 | 3400930024218 |
| NICOPATCHLIB | 7 mg/24h | 7 | 3400930024164 |
| NICOPATCH | 21 mg/24h | 28 | 3400938381009 |
| NICOPATCH | 21 mg/24h | 7 | 3400938380637 |
| NICOPATCH | 14 mg/24h | 28 | 3400938380576 |
| NICOPATCH | 14 mg/24h | 7 | 3400938380286 |
| NICOPATCH | 7 mg/24h | 28 | 3400938380118 |
| NICOPATCH | 7 mg/24h | 7 | 3400938379808 |
| NICORETTESKIN | 25 mg/16h | 28 | 3400939651231 |
| NICORETTESKIN | 25 mg/16h | 7 | 3400939651002 |
| NICORETTESKIN | 15 mg/16h | 28 | 3400939650920 |
| NICORETTESKIN | 15 mg/16h | 7 | 3400939650630 |
| NICORETTESKIN | 10 mg/16h | 28 | 3400939650579 |
| NICORETTESKIN | 10 mg/16h | 7 | 3400939650340 |
| NIQUITIN | 21 mg/24h | 28 | 3400937955959 |
| NIQUITIN | 21 mg/24h | 7 | 3400937955720 |
| NIQUITIN | 14 mg/24h | 28 | 3400937956321 |
| NIQUITIN | 14 mg/24h | 7 | 3400937956031 |
| NIQUITIN | 7 mg/24h | 28 | 3400935719676 |
| NIQUITIN | 7 mg/24h | 7 | 3400937956499 |
| NICOTINELL TTS | 21 mg/24h | 28 | 3400933468743 |
| NICOTINELL TTS | 21 mg/24h | 7 | 3400933468453 |
| NICOTINELL TTS | 14 mg/24h | 28 | 3400933468392 |
| NICOTINELL TTS | 14 mg/24h | 7 | 3400933467913 |
| NICOTINELL TTS | 7 mg/24h | 28 | 3400933467852 |
| NICOTINELL TTS | 7 mg/24h | 7 | 3400933467562 |
| **Non-patch form** |  |  |  |
| **Gum** |  |  |  |
| NICORETTE FRUITS SANS SUCRE | 4 mg | 105 | 3400937100731 |
| NICORETTE FRUITS SANS SUCRE | 2 mg | 210 | 3400937576956 |
| NICORETTE FRUITS SANS SUCRE | 2 mg | 105 | 3400937099578 |
| NICORETTE SANS SUCRE | 4 mg | 105 | 3400937630757 |
| NICORETTE SANS SUCRE | 2 mg | 210 | 3400937631297 |
| NICORETTE SANS SUCRE | 2 mg | 105 | 3400937631129 |
| NICORETTE MENTHE FRAICHE SANS SUCRE | 4 mg | 105 | 3400937631587 |
| NICORETTE MENTHE FRAICHE SANS SUCRE | 2 mg | 210 | 3400937631419 |
| NICORETTE MENTHE FRAICHE SANS SUCRE | 2 mg | 105 | 3400937631358 |
| NICORETTE MENTHE GLACIALE SANS SUCRE | 4 mg | 105 | 3400937098397 |
| NICORETTE MENTHE GLACIALE SANS SUCRE | 2 mg | 210 | 3400939003368 |
| NICORETTE MENTHE GLACIALE SANS SUCRE | 2 mg | 105 | 3400937096966 |
| NICOTINE EG FRUIT | 4 mg | 108 | 3400926896492 |
| NICOTINE EG FRUIT | 2 mg | 108 | 3400926893071 |
| NICOTINE EG MENTHE | 4 mg | 108 | 3400926903169 |
| NICOTINE EG MENTHE | 2 mg | 108 | 3400926899745 |
| NIQUITIN MENTHE GLACIALE | 4 mg | 100 | 3400930096796 |
| NIQUITIN MENTHE GLACIALE | 4 mg | 30 | 3400930096789 |
| NIQUITIN MENTHE GLACIALE | 2 mg | 100 | 3400930096635 |
| NIQUITIN MENTHE GLACIALE | 2 mg | 30 | 3400930096628 |
| NIQUITIN FRUITS TROPICAUX | 4 mg | 100 | 3400930096598 |
| NIQUITIN FRUITS TROPICAUX | 4 mg | 30 | 3400930096581 |
| NIQUITIN FRUITS TROPICAUX | 2 mg | 100 | 3400930096499 |
| NIQUITIN FRUITS TROPICAUX | 2 mg | 30 | 3400930096482 |
| NICOTINELL FRUIT | 2 mg | 204 | 3400939395227 |
| NICOTINELL FRUIT | 2 mg | 96 | 3400936317123 |
| NICOTINELL FRUIT | 2 mg | 36 | 3400936316751 |
| NICOTINELL FRUIT EXOTIQUE | 2 mg | 204 | 3400921796124 |
| NICOTINELL FRUIT EXOTIQUE | 2 mg | 96 | 3400921796063 |
| NICOTINELL MENTHE FRAICHEUR | 4 mg | 96 | 3400936316232 |
| NICOTINELL MENTHE FRAICHEUR | 4 mg | 36 | 3400936315860 |
| NICOTINELL MENTHE FRAICHEUR | 2 mg | 204 | 3400939395456 |
| NICOTINELL MENTHE FRAICHEUR | 2 mg | 96 | 3400936315341 |
| NICOTINELL MENTHE FRAICHEUR | 2 mg | 36 | 3400936314979 |
| Tablets |  |  |  |
| NIQUITIN | 4 mg | 96 | 3400934702785 |
| NIQUITIN | 2 mg | 96 | 3400934702846 |
| NIQUITIN | 2 mg | 36 | 3400935752383 |
| NIQUITINMINIS | 4 mg | 60 | 3400938672770 |
| NIQUITINMINIS | 4 mg | 20 | 3400938672541 |
| NIQUITINMINIS | 1,5 mg | 60 | 3400938672312 |
| NIQUITINMINIS | 1,5 mg | 20 | 3400938672190 |
| NICOTINELL | 2 mg | 204 | 3400936868601 |
| NICOTINELL | 2 mg | 144 | 3400938924473 |
| NICOTINELL | 2 mg | 96 | 3400936868540 |
| NICOTINELL | 2 mg | 36 | 3400936868489 |
| NICOTINELL | 1 mg | 204 | 3400936409125 |
| NICOTINELL | 1 mg | 144 | 3400938919912 |
| NICOTINELL | 1 mg | 96 | 3400936409064 |
| NICOTINELL | 1 mg | 36 | 3400936408982 |
| NICORETTE MICROTAB | 2 mg | 150 | 3400937992459 |
| NICORETTE MICROTAB | 2 mg | 100 | 3400937992398 |
| NICORETTE MICROTAB CITRON | 2 mg | 100 | 3400937986946 |
| Lozenge |  |  |  |
| NICOPASS SANS SUCRE EUCALYPTUS | 1,5 mg | 96 | 3400937680554 |
| NICOPASS SANS SUCRE MENTHE FRAICHEUR | 1,5 mg | 96 | 3400936498990 |
| NICOPASS SANS SUCRE REGLISSE | 1,5 mg | 96 | 3400936499881 |
| NICOPASS MENTHE FRAICHEUR | 2,5 mg | 96 | 3400938730715 |
| NICOPASS REGLISSE MENTHE | 2,5 mg | 96 | 3400938729535 |

**Table S2.** Definition of exclusion criteria.

| Codes |  |
| --- | --- |
| **Cardiovascular disease** |  |
| Heart failure |  |
| ICD-10 codes | I50, J81, I11, I13, K761 |
| Ischemic heart disease |  |
| ICD-10 codes | I20-I25 |
| Arrhythmia |  |
| ICD-10 codes | I44-I45, I47-I49 |
| Stroke |  |
| ICD-10 codes | G45-G46, I60-I69 |
| **Respiratory disease** |  |
| COPD/asthma |  |
| ICD-10 codes | J40-J42, J44, J961+0 |
| Bronchodilators |  |
| ATC code | R03 |
| **Cancer** |  |
| ICD-10 codes | C00-C75, C77-C80, D00-D09, D37-D48, Z51.0, Z51.1 |

ATC Anatomic therapeutic chemical; COPD Chronic obstructive pulmonary disease; ICD-10 International classification of diseases, 10<sup>th</sup> edition; LTD Long-Term Disease

**Table S3.** Definition of covariates.

| **Codes** |  |
| --- | --- |
| **Lifestyle habits** |  |
| **Alcohol use disorders** |  |
| ICD-10 codes | E24.4, E51.2, F10, G31.2, G62.1, G72.1, I42.6, K29.2, K70, K86.0, R78.0, T51, X45, X65, Y15, Y90, Y91, Y57.3, Z50.2, Z71.4, Z72.1 |
| ATC codes | N07BB01, N07BB03, N07BB04, N07BB05 |
| ATC and ICD-10 codes | M03BX01 and no ICD-10 codes in C70, C71, C79.3, C79.4, D32, D33, D42, D43, G04-G06, G09, G12, G13, G24-26, G31, G32, G35-G37, G46, G80-G83, G91, G93, G95 |
| **Smoking use disorders** |  |
| ICD-10 codes | Z71.6, Z72.0, F17, T65.2 |
| ATC codes | N06AX12, N07BA |
| Healthcare delivery codes* | 9566, 9526, 9527 |
| **Substance abuse** |  |
| ICD-10 codes | F11-F16, F18-F19, Z71.5, Z72.2 |
| ATC codes | N07BC |
| **Comorbidities** |  |
| **Morbid obesity** |  |
| ICD-10 codes | E65 – E68 |
| **Diabetes** |  |
| ICD-10 codes | E10 - E14 |
|  | At least one ICD-10 in E10-E14 (associated diagnostic) and at least one ICD-10 in G59.0, G63.2, G99.0, H28.0, H36.0, I79.2, L97, M14.2, M14.6, N08.3 |
| ATC codes | A10AB01, A10AB03, A10AB04, A10AB05, A10AB06, A10AC01, A10AC03, A10AC04, A10AD01, A10AD03, A10AD04, A10AD05, A10AE01, A10AE02, A10AE03, A10AE04, A10AE05, A10AE30, A10BA02, A10BB01, A10BB03, A10BB04, A10BB06, A10BB07, A10BB09, A10BB12, A10BD02, A10BD03, A10BD05, A10BD07, A10BD08, A10BD10, A10BD15, A10BD16, A10BF01, A10BF02, A10BG02, A10BG03, A10BH01, A10BH02, A10BH03, A10BX02, A10BX04, A10BX07, A10BX09, A10BX10, A10BX11, A10BX12 |
| **Dementia** |  |
| ICD-10 codes | F00 - F03, F051, G30 |
| **Suicide attempt** |  |
| ICD-10 codes | X60 - X84 |

| Medication : ATC codes |  |
| --- | --- |
| Antihypertensive drugs |  |
| ACE inhibitor | C09AA, C09BA01, C09BA02, C09BA03, C09BA04, C09BA05, C09BA06, C09BA07, C09BA09, C09BA15 |
| ATC codes | C02AB02, C02AC01, C02AC02, C02AC05, C02AC06, C02CA01, C02CA06, C02DC01, C02LA01, C03AA01, C03AA03, C03BA04, C03BA10, C03BA11, C03BX03, C03CA01, C03CA02, C03CA03, C03DA01, C03DB01, C03EA, C03EA01, C03EA04, C07AA02, C07AA03, C07AA05, C07AA06, C07AA12, C07AA15, C07AA16, C07AA23, C07AB02, C07AB03, C07AB04, C07AB05, C07AB07, C07AB08, C07AB12, C07AG01, C07AG02, C07BA02, C07BB02, C07BB03, C07BB07, C07BB12, C07CA03, C07DA06, C07FB02, C07FB03, C08CA01, C08CA02, C08CA03, C08CA04, C08CA05, C08CA06, C08CA08, C08CA09, C08CA11, C08CA13, C08CX01, C08DA01, C08DB01, C08EA02, C08GA02, C09BB02, C09BB04, C09CA01, C09CA02, C09CA03, C09CA04, C09CA06, C09CA07, C09CA08, C09DA01, C09DA02, C09DA03, C09DA04, C09DA06, C09DA07, C09DA08, C09DB01, C09DB02, C09DB04, C09XA02, C09XA52 |
| Lipid lowering drugs | C10AA01, C10AA03, C10AA04, C10AA05, C10AA06, C10AA07, C10AB01, C10AB02, C10AB04, C10AB05, C10AB08, C10AX09, C10BA02, C10BX02, C10BX03 |
| Psychiatric drugs |  |
| Antidepressants | N06A |
| Anxiolytics | N05BA01, N05BA04, N05BA05, N05BA06, N05BA08, N05BA09, N05BA11, N05BA12, N05BA16, N05BA18, N05BA21, N05BA23, N05BB01, N05BB02, N05BC01, N05BE01, N05BX03 |
| Hypnotics | N05BC51, N05CD02, N05CD03, N05CD04, N05CD05, N05CD06, N05CD07, N05CD11, N05CF01, N05CF02, N05CM11, N05CM16, N05CX |
| Neuroleptics | N05A except N05AN, N05AN01 and N05AL06 |
| NSAID | M01AE01, M01AE02, M01AE03, M01AE09, M01AE11, M01AE16, M01AB01, M01AB05, M01AB08, M01AB16, M01AC01, M01AC02, M01AC06, M01AH01, M01AH05, M01AX01, M01AX02, M01AX17, M01AX21, M01AX22 |
| Oral corticosteroids | H02AB01, H02AB02, H02AB04, H02AB06, H02AB07, H02AB09, H02AB10 |

ACE Angiotensin converting enzyme; ATC Anatomical Therapeutic Chemical; ICD-10 International classification of diseases, 10<sup>th</sup> edition

**Table S4.** Odds ratios (95% CIs) of exposure to nicotine replacement therapy for all variables included in the propensity score model (logistic regression) on population without smoking-related diseases.

| Characteristic* | OR (CI 95%) |
| --- | --- |
| Female sex | 1.00 (0.99 – 1.01) |
| Age (/10 years old) | 0.92 (0.92 – 0.93) |
| Regions |  |
| Normandie | Ref |
| Ile de France<sup>§</sup> | 1.08 (1.05 – 1.11) |
| Centre-Val de Loire | 1.02 (0.99 – 1.05) |
| Bourgogne Franche Comté | 0.99 (0.96 – 1.02) |
| Hauts de France | 1.00 (0.98 – 1.02) |
| Grand Est | 1.04 (1.02 – 1.07) |
| Pays de Loire | 1.03 (1.01 – 1.06) |
| Bretagne | 0.99 (0.96 – 1.02) |
| Nouvelle Aquitaine | 1.03 (1.00 – 1.05) |
| Occitanie | 1.04 (1.02 – 1.07) |
| Auvergne-Rhône Alpes | 1.04 (1.01 – 1.06) |
| Provence Alpes Cote | 1.04 (1.01 – 1.06) |
| Corse | 1.25 (1.16 – 1.36) |
| CMU_C | 1.20 (1.19 – 1.22) |
| Social deprivation index (quintiles) |  |
| 1 (the least deprivation) | Ref |
| 2 | 1.06 (1.05 – 1.08) |
| 3 | 1.08 (1.07 – 1.10) |
| 4 | 1.10 (1.08 – 1.12) |
| 5 (the most deprivation) | 1.09 (1.08 – 1.11) |
| Obesity | 1.63 (1.57 – 1.69) |
| Diabetes | 0.92 (0.90 – 0.95) |
| Dementia | 0.51 (0.41 – 0.62) |
| Alcohol use disorders | 3.38 (3.28 – 3.47) |
| Substance abuse | 2.82 (2.71 – 2.94) |
| Suicide attempt | 1.26 (1.20 – 1.33) |
| Anti-hypertensive drugs<sup>a</sup> |  |
| No | Ref |
| ACE | 1.14 (1.12 – 1.17) |
| others | 0.99 (0.97 – 1.00) |
| Lipid lowering drugs<sup>a</sup> | 1.70 (1.67 – 1.73) |
| Antidepressants<sup>a</sup> | 1.58 (1.56 – 1.61) |
| Anxiolytics<sup>a</sup> | 1.87 (1.84 – 1.90) |
| Hypnotics<sup>a</sup> | 1.47 (1.43 – 1.52) |
| Neuroleptics<sup>a</sup> | 2.02 (1.97 – 2.08) |
| NSAID<sup>a</sup> | 1.49 (1.46 – 1.51) |
| Oral Corticosteroids<sup>a</sup> | 1.62 (1.56 – 1.67) |

\* CMU\_C Complementary Universal health insurance; ACE Angiotensin-converting enzyme; NSAID Non-steroidal anti-inflammatory drug
<sup>§</sup>Paris and great Paris
<sup>a</sup>At least three dispensations in previous year

**Table S5.** Odds ratios (95% CIs) of exposure to nicotine replacement therapy for all variables included in the propensity score model (logistic regression) on population with exposed individual with smoking-related diseases.

| Characteristic* | OR (CI 95%) |
| --- | --- |
| Female sex | 1.09 (1.07 – 1.11) |
| Age (/10 years old) | 0.80 (0.79 – 0.81) |
| Regions |  |
| Normandie | Ref |
| Ile de France<sup>§</sup> | 1.23 (1.18 – 1.28) |
| Centre-Val de Loire | 1.05 (1.00 – 1.10) |
| Bourgogne Franche Comté | 1.02 (0.97 – 1.07) |
| Hauts de France | 0.94 (0.90 – 0.98) |
| Grand Est | 1.06 (1.02 – 1.10) |
| Pays de Loire | 1.12 (1.07 – 1.17) |
| Bretagne | 1.03 (0.99 – 1.08) |
| Nouvelle Aquitaine | 1.08 (1.04 – 1.12) |
| Occitanie | 1.14 (1.10 – 1.19) |
| Auvergne-Rhône Alpes | 1.13 (1.08 – 1.17) |
| Provence Alpes Cote | 1.16 (1.11 – 1.21) |
| Corse | 1.68 (1.46 – 1.92) |
| CMU_C | 1.31 (1.28 – 1.34) |
| Social deprivation index (quintiles) |  |
| 1 (the least deprivation) | Ref |
| 2 | 1.08 (1.05 – 1.11) |
| 3 | 1.11 (1.08 – 1.14) |
| 4 | 1.13 (1.10 – 1.16) |
| 5 (the most deprivation) | 1.16 (1.13 – 1.19) |
| Unknown | 0.79 (0.72 – 0.85) |
| Obesity | 1.40 (1.37 – 1.44) |
| Diabetes | 0.82 (0.80 – 0.84) |
| Dementia | 0.39 (0.33 – 0.46) |
| Alcohol use disorders | 5.91 (5.70 – 6.13) |
| Substance abuse | 4.05 (3.80 – 4.32) |
| Suicide attempt | 1.36 (1.26 – 1.48) |
| Anti-hypertensive drugs<sup>a</sup> |  |
| No | Ref |
| ACE | 2.34 (2.28 – 2.41) |
| others | 1.45 (1.42 – 1.48) |
| Lipid lowering drugs<sup>a</sup> | 2.66 (2.60 – 2.72) |
| Antidepressants<sup>a</sup> | 1.61 (1.57 – 1.65) |
| Anxiolytics<sup>a</sup> | 2.34 (2.28 – 2.40) |
| Hypnotics<sup>a</sup> | 1.84 (1.78 – 1.90) |
| Neuroleptics<sup>a</sup> | 1.76 (1.68 – 1.83) |
| NSAID<sup>a</sup> | 1.18 (1.16 – 1.21) |
| Oral Corticosteroids<sup>a</sup> | 4.20 (4.06 – 4.35) |

\*CMU\_C Complementary Universal health insurance; ACE Angiotensin-converting enzyme; NSAID Non-steroidal anti-inflammatory drug
<sup>§</sup>Paris and great Paris
<sup>a</sup>At least three dispensations in previous year

**Table S6.**
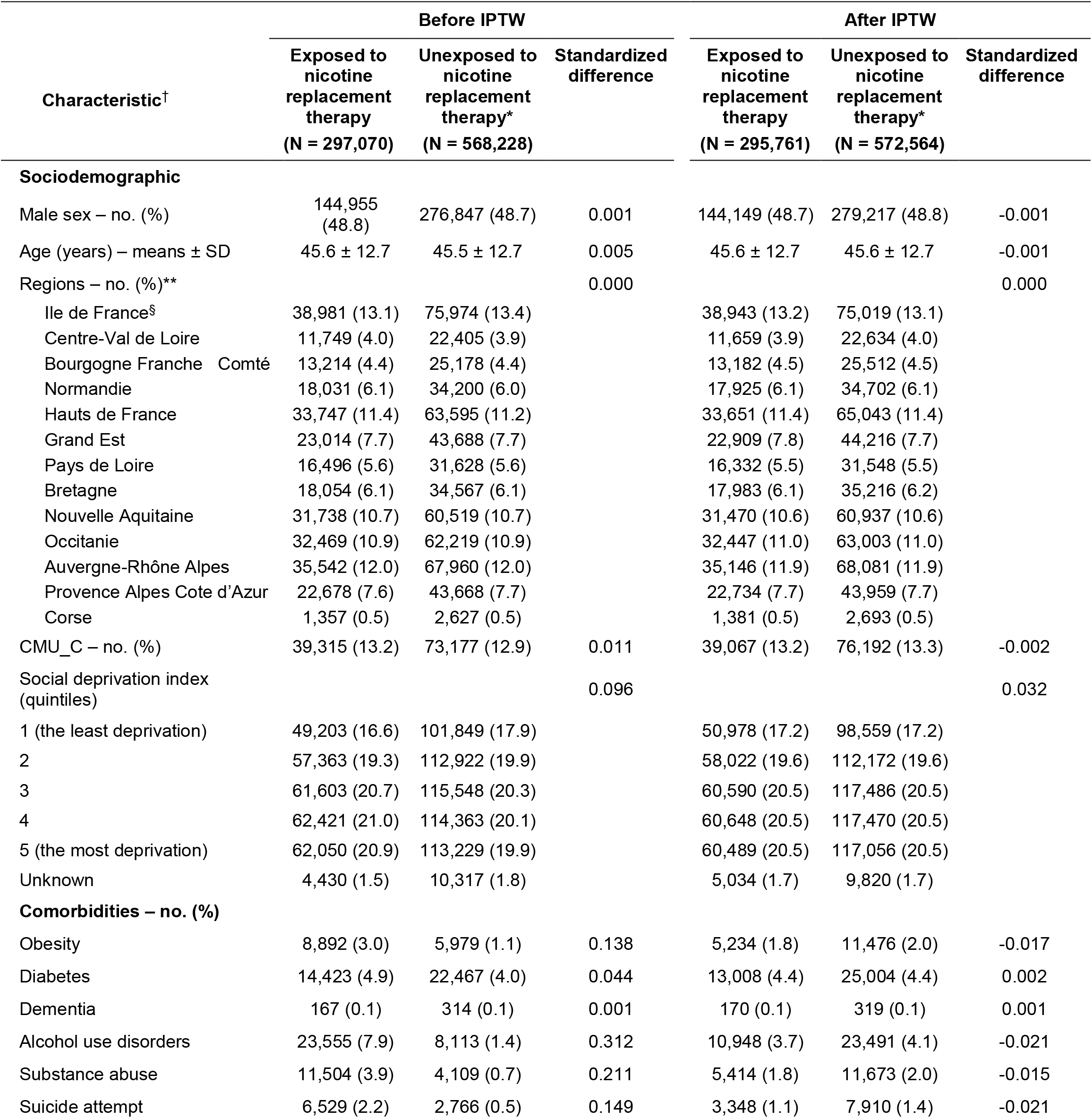
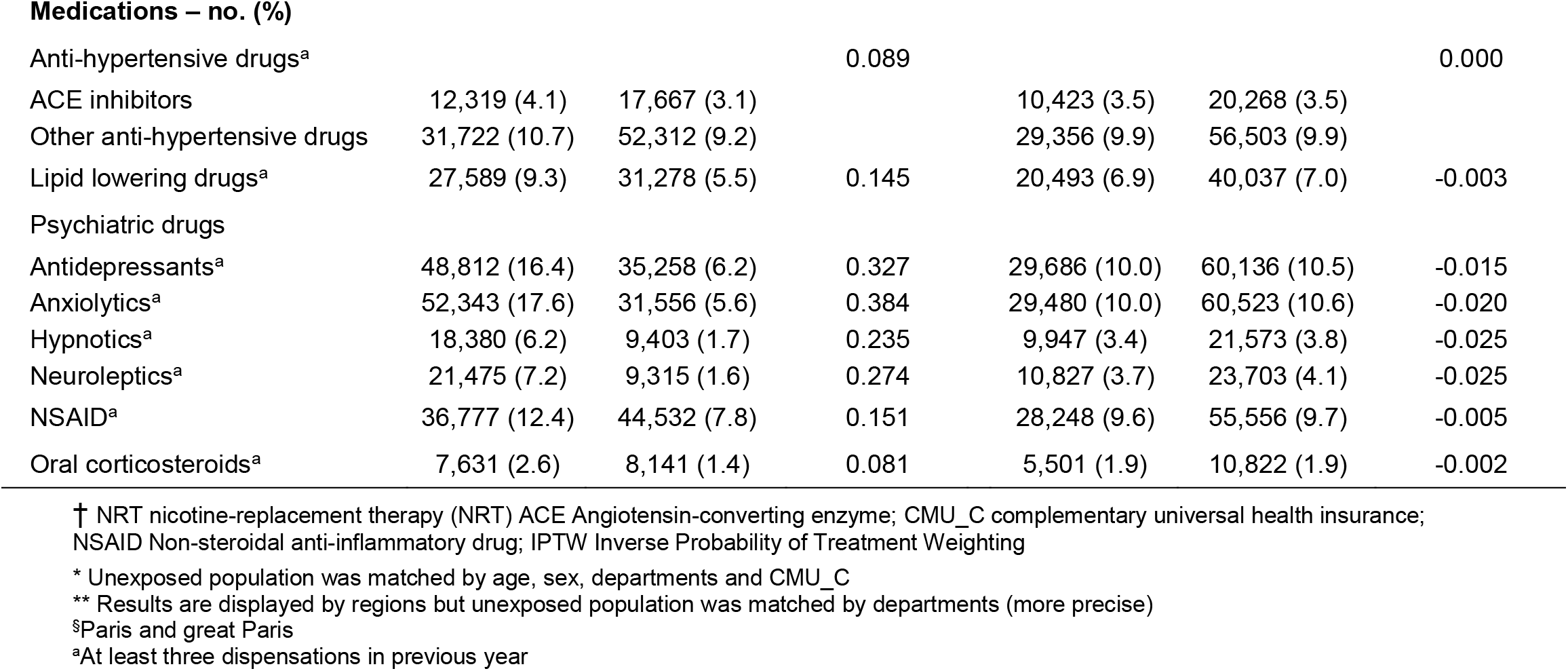
Baseline characteristics of participants according to nicotine-replacement therapy (NRT) exposure before and after inverse probability of treatment weighting (IPTW) on population without smoking-related diseases.

| Characteristic<sup>†</sup> | Before IPTW |  |  | After IPTW |  |  |
| --- | --- | --- | --- | --- | --- | --- |
|  | Exposed to nicotine replacement therapy<br>(N = 297,070) | Unexposed to nicotine replacement therapy*<br>(N = 568,228) | Standardized difference | Exposed to nicotine replacement therapy<br>(N = 295,761) | Unexposed to nicotine replacement therapy*<br>(N = 572,564) | Standardized difference |
| **Sociodemographic** |  |  |  |  |  |  |
| Male sex – no. (%) | 144,955 (48.8) | 276,847 (48.7) | 0.001 | 144,149 (48.7) | 279,217 (48.8) | -0.001 |
| Age (years) – means ± SD | 45.6 ± 12.7 | 45.5 ± 12.7 | 0.005 | 45.6 ± 12.7 | 45.6 ± 12.7 | -0.001 |
| Regions – no. (%)** |  |  | 0.000 |  |  | 0.000 |
| Ile de France<sup>§</sup> | 38,981 (13.1) | 75,974 (13.4) |  | 38,943 (13.2) | 75,019 (13.1) |  |
| Centre-Val de Loire | 11,749 (4.0) | 22,405 (3.9) |  | 11,659 (3.9) | 22,634 (4.0) |  |
| Bourgogne Franche Comté | 13,214 (4.4) | 25,178 (4.4) |  | 13,182 (4.5) | 25,512 (4.5) |  |
| Normandie | 18,031 (6.1) | 34,200 (6.0) |  | 17,925 (6.1) | 34,702 (6.1) |  |
| Hauts de France | 33,747 (11.4) | 63,595 (11.2) |  | 33,651 (11.4) | 65,043 (11.4) |  |
| Grand Est | 23,014 (7.7) | 43,688 (7.7) |  | 22,909 (7.8) | 44,216 (7.7) |  |
| Pays de Loire | 16,496 (5.6) | 31,628 (5.6) |  | 16,332 (5.5) | 31,548 (5.5) |  |
| Bretagne | 18,054 (6.1) | 34,567 (6.1) |  | 17,983 (6.1) | 35,216 (6.2) |  |
| Nouvelle Aquitaine | 31,738 (10.7) | 60,519 (10.7) |  | 31,470 (10.6) | 60,937 (10.6) |  |
| Occitanie | 32,469 (10.9) | 62,219 (10.9) |  | 32,447 (11.0) | 63,003 (11.0) |  |
| Auvergne-Rhône Alpes | 35,542 (12.0) | 67,960 (12.0) |  | 35,146 (11.9) | 68,081 (11.9) |  |
| Provence Alpes Cote d'Azur | 22,678 (7.6) | 43,668 (7.7) |  | 22,734 (7.7) | 43,959 (7.7) |  |
| Corse | 1,357 (0.5) | 2,627 (0.5) |  | 1,381 (0.5) | 2,693 (0.5) |  |
| CMU_C – no. (%) | 39,315 (13.2) | 73,177 (12.9) | 0.011 | 39,067 (13.2) | 76,192 (13.3) | -0.002 |
| Social deprivation index (quintiles) |  |  | 0.096 |  |  | 0.032 |
| 1 (the least deprivation) | 49,203 (16.6) | 101,849 (17.9) |  | 50,978 (17.2) | 98,559 (17.2) |  |
| 2 | 57,363 (19.3) | 112,922 (19.9) |  | 58,022 (19.6) | 112,172 (19.6) |  |
| 3 | 61,603 (20.7) | 115,548 (20.3) |  | 60,590 (20.5) | 117,486 (20.5) |  |
| 4 | 62,421 (21.0) | 114,363 (20.1) |  | 60,648 (20.5) | 117,470 (20.5) |  |
| 5 (the most deprivation) | 62,050 (20.9) | 113,229 (19.9) |  | 60,489 (20.5) | 117,056 (20.5) |  |
| Unknown | 4,430 (1.5) | 10,317 (1.8) |  | 5,034 (1.7) | 9,820 (1.7) |  |
| **Comorbidities – no. (%)** |  |  |  |  |  |  |
| Obesity | 8,892 (3.0) | 5,979 (1.1) | 0.138 | 5,234 (1.8) | 11,476 (2.0) | -0.017 |
| Diabetes | 14,423 (4.9) | 22,467 (4.0) | 0.044 | 13,008 (4.4) | 25,004 (4.4) | 0.002 |
| Dementia | 167 (0.1) | 314 (0.1) | 0.001 | 170 (0.1) | 319 (0.1) | 0.001 |
| Alcohol use disorders | 23,555 (7.9) | 8,113 (1.4) | 0.312 | 10,948 (3.7) | 23,491 (4.1) | -0.021 |
| Substance abuse | 11,504 (3.9) | 4,109 (0.7) | 0.211 | 5,414 (1.8) | 11,673 (2.0) | -0.015 |
| Suicide attempt | 6,529 (2.2) | 2,766 (0.5) | 0.149 | 3,348 (1.1) | 7,910 (1.4) | -0.021 |
| Medications – no. (%) |  |  |  |  |  |  |
| Anti-hypertensive drugs<sup>a</sup> |  |  | 0.089 |  |  | 0.000 |
| ACE inhibitors | 12,319 (4.1) | 17,667 (3.1) |  | 10,423 (3.5) | 20,268 (3.5) |  |
| Other anti-hypertensive drugs | 31,722 (10.7) | 52,312 (9.2) |  | 29,356 (9.9) | 56,503 (9.9) |  |
| Lipid lowering drugs<sup>a</sup> | 27,589 (9.3) | 31,278 (5.5) | 0.145 | 20,493 (6.9) | 40,037 (7.0) | -0.003 |
| Psychiatric drugs |  |  |  |  |  |  |
| Antidepressants<sup>a</sup> | 48,812 (16.4) | 35,258 (6.2) | 0.327 | 29,686 (10.0) | 60,136 (10.5) | -0.015 |
| Anxiolytics<sup>a</sup> | 52,343 (17.6) | 31,556 (5.6) | 0.384 | 29,480 (10.0) | 60,523 (10.6) | -0.020 |
| Hypnotics<sup>a</sup> | 18,380 (6.2) | 9,403 (1.7) | 0.235 | 9,947 (3.4) | 21,573 (3.8) | -0.025 |
| Neuroleptics<sup>a</sup> | 21,475 (7.2) | 9,315 (1.6) | 0.274 | 10,827 (3.7) | 23,703 (4.1) | -0.025 |
| NSAID<sup>a</sup> | 36,777 (12.4) | 44,532 (7.8) | 0.151 | 28,248 (9.6) | 55,556 (9.7) | -0.005 |
| Oral corticosteroids<sup>a</sup> | 7,631 (2.6) | 8,141 (1.4) | 0.081 | 5,501 (1.9) | 10,822 (1.9) | -0.002 |

† NRT nicotine-replacement therapy (NRT) ACE Angiotensin-converting enzyme; CMU\_C complementary universal health insurance; NSAID Non-steroidal anti-inflammatory drug; IPTW Inverse Probability of Treatment Weighting
\* Unexposed population was matched by age, sex, departments and CMU\_C
\*\* Results are displayed by regions but unexposed population was matched by departments (more precise)
§Paris and great Paris
<sup>a</sup>At least three dispensations in previous year

**Table S7.**
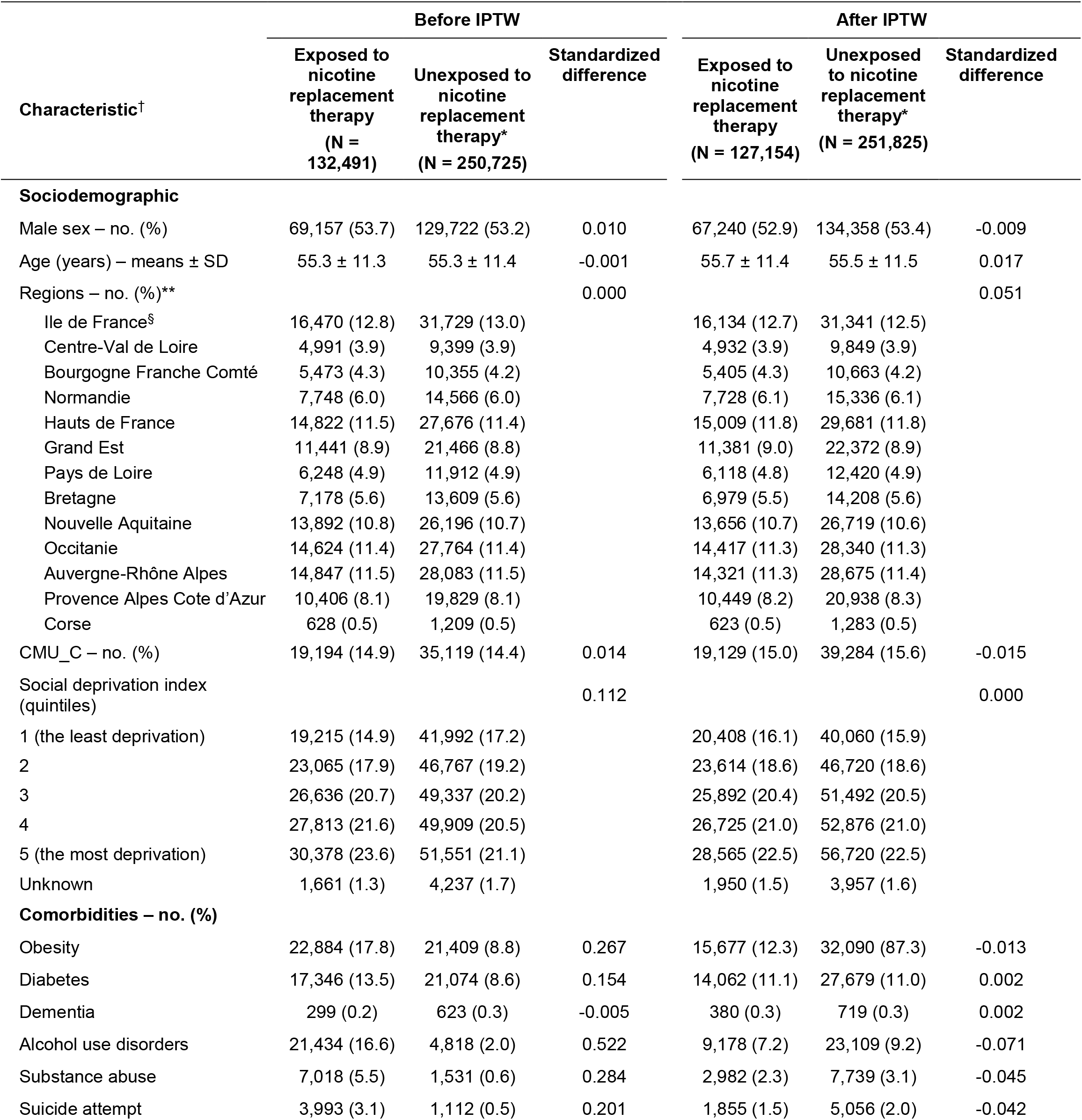
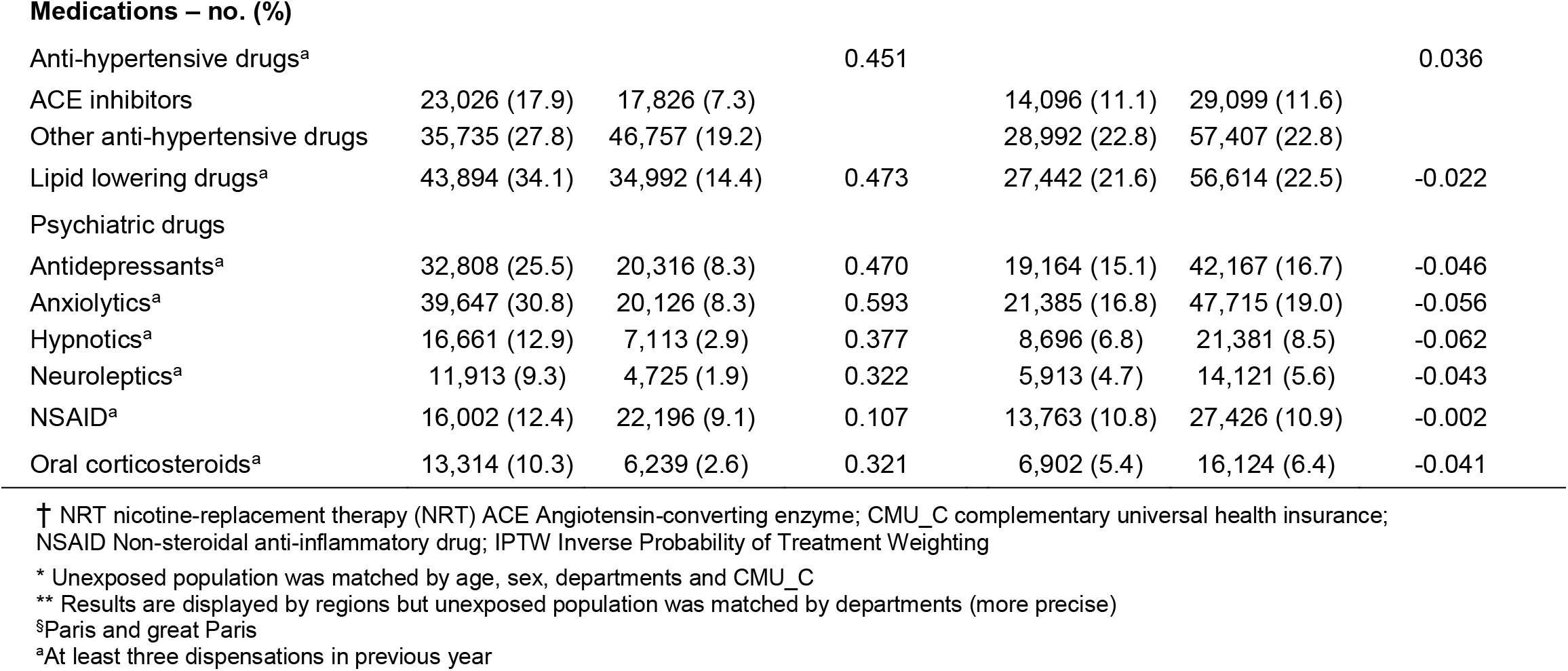
Baseline characteristics of participants according to nicotine-replacement therapy (NRT) exposure before and after inverse probability of treatment weighting (IPTW) on population with exposed individual with smoking-related diseases.

| Characteristic<sup>†</sup> | Before IPTW |  |  | After IPTW |  |  |
| --- | --- | --- | --- | --- | --- | --- |
|  | Exposed to nicotine replacement therapy<br>(N = 132,491) | Unexposed to nicotine replacement therapy*<br>(N = 250,725) | Standardized difference | Exposed to nicotine replacement therapy<br>(N = 127,154) | Unexposed to nicotine replacement therapy*<br>(N = 251,825) | Standardized difference |
| **Sociodemographic** |  |  |  |  |  |  |
| Male sex – no. (%) | 69,157 (53.7) | 129,722 (53.2) | 0.010 | 67,240 (52.9) | 134,358 (53.4) | -0.009 |
| Age (years) – means ± SD | 55.3 ± 11.3 | 55.3 ± 11.4 | -0.001 | 55.7 ± 11.4 | 55.5 ± 11.5 | 0.017 |
| Regions – no. (%)** |  |  | 0.000 |  |  | 0.051 |
| Ile de France<sup>§</sup> | 16,470 (12.8) | 31,729 (13.0) |  | 16,134 (12.7) | 31,341 (12.5) |  |
| Centre-Val de Loire | 4,991 (3.9) | 9,399 (3.9) |  | 4,932 (3.9) | 9,849 (3.9) |  |
| Bourgogne Franche Comté | 5,473 (4.3) | 10,355 (4.2) |  | 5,405 (4.3) | 10,663 (4.2) |  |
| Normandie | 7,748 (6.0) | 14,566 (6.0) |  | 7,728 (6.1) | 15,336 (6.1) |  |
| Hauts de France | 14,822 (11.5) | 27,676 (11.4) |  | 15,009 (11.8) | 29,681 (11.8) |  |
| Grand Est | 11,441 (8.9) | 21,466 (8.8) |  | 11,381 (9.0) | 22,372 (8.9) |  |
| Pays de Loire | 6,248 (4.9) | 11,912 (4.9) |  | 6,118 (4.8) | 12,420 (4.9) |  |
| Bretagne | 7,178 (5.6) | 13,609 (5.6) |  | 6,979 (5.5) | 14,208 (5.6) |  |
| Nouvelle Aquitaine | 13,892 (10.8) | 26,196 (10.7) |  | 13,656 (10.7) | 26,719 (10.6) |  |
| Occitanie | 14,624 (11.4) | 27,764 (11.4) |  | 14,417 (11.3) | 28,340 (11.3) |  |
| Auvergne-Rhône Alpes | 14,847 (11.5) | 28,083 (11.5) |  | 14,321 (11.3) | 28,675 (11.4) |  |
| Provence Alpes Cote d’Azur | 10,406 (8.1) | 19,829 (8.1) |  | 10,449 (8.2) | 20,938 (8.3) |  |
| Corse | 628 (0.5) | 1,209 (0.5) |  | 623 (0.5) | 1,283 (0.5) |  |
| CMU_C – no. (%) | 19,194 (14.9) | 35,119 (14.4) | 0.014 | 19,129 (15.0) | 39,284 (15.6) | -0.015 |
| Social deprivation index (quintiles) |  |  | 0.112 |  |  | 0.000 |
| 1 (the least deprivation) | 19,215 (14.9) | 41,992 (17.2) |  | 20,408 (16.1) | 40,060 (15.9) |  |
| 2 | 23,065 (17.9) | 46,767 (19.2) |  | 23,614 (18.6) | 46,720 (18.6) |  |
| 3 | 26,636 (20.7) | 49,337 (20.2) |  | 25,892 (20.4) | 51,492 (20.5) |  |
| 4 | 27,813 (21.6) | 49,909 (20.5) |  | 26,725 (21.0) | 52,876 (21.0) |  |
| 5 (the most deprivation) | 30,378 (23.6) | 51,551 (21.1) |  | 28,565 (22.5) | 56,720 (22.5) |  |
| Unknown | 1,661 (1.3) | 4,237 (1.7) |  | 1,950 (1.5) | 3,957 (1.6) |  |
| **Comorbidities – no. (%)** |  |  |  |  |  |  |
| Obesity | 22,884 (17.8) | 21,409 (8.8) | 0.267 | 15,677 (12.3) | 32,090 (87.3) | -0.013 |
| Diabetes | 17,346 (13.5) | 21,074 (8.6) | 0.154 | 14,062 (11.1) | 27,679 (11.0) | 0.002 |
| Dementia | 299 (0.2) | 623 (0.3) | -0.005 | 380 (0.3) | 719 (0.3) | 0.002 |
| Alcohol use disorders | 21,434 (16.6) | 4,818 (2.0) | 0.522 | 9,178 (7.2) | 23,109 (9.2) | -0.071 |
| Substance abuse | 7,018 (5.5) | 1,531 (0.6) | 0.284 | 2,982 (2.3) | 7,739 (3.1) | -0.045 |
| Suicide attempt | 3,993 (3.1) | 1,112 (0.5) | 0.201 | 1,855 (1.5) | 5,056 (2.0) | -0.042 |
| Medications – no. (%) |  |  |  |  |  |  |
| Anti-hypertensive drugs<sup>a</sup> |  |  | 0.451 |  |  | 0.036 |
| ACE inhibitors | 23,026 (17.9) | 17,826 (7.3) |  | 14,096 (11.1) | 29,099 (11.6) |  |
| Other anti-hypertensive drugs | 35,735 (27.8) | 46,757 (19.2) |  | 28,992 (22.8) | 57,407 (22.8) |  |
| Lipid lowering drugs<sup>a</sup> | 43,894 (34.1) | 34,992 (14.4) | 0.473 | 27,442 (21.6) | 56,614 (22.5) | -0.022 |
| Psychiatric drugs |  |  |  |  |  |  |
| Antidepressants<sup>a</sup> | 32,808 (25.5) | 20,316 (8.3) | 0.470 | 19,164 (15.1) | 42,167 (16.7) | -0.046 |
| Anxiolytics<sup>a</sup> | 39,647 (30.8) | 20,126 (8.3) | 0.593 | 21,385 (16.8) | 47,715 (19.0) | -0.056 |
| Hypnotics<sup>a</sup> | 16,661 (12.9) | 7,113 (2.9) | 0.377 | 8,696 (6.8) | 21,381 (8.5) | -0.062 |
| Neuroleptics<sup>a</sup> | 11,913 (9.3) | 4,725 (1.9) | 0.322 | 5,913 (4.7) | 14,121 (5.6) | -0.043 |
| NSAID<sup>a</sup> | 16,002 (12.4) | 22,196 (9.1) | 0.107 | 13,763 (10.8) | 27,426 (10.9) | -0.002 |
| Oral corticosteroids<sup>a</sup> | 13,314 (10.3) | 6,239 (2.6) | 0.321 | 6,902 (5.4) | 16,124 (6.4) | -0.041 |

† NRT nicotine-replacement therapy (NRT) ACE Angiotensin-converting enzyme; CMU\_C complementary universal health insurance; NSAID Non-steroidal anti-inflammatory drug; IPTW Inverse Probability of Treatment Weighting
\* Unexposed population was matched by age, sex, departments and CMU\_C
\*\* Results are displayed by regions but unexposed population was matched by departments (more precise)
§Paris and great Paris
<sup>a</sup>At least three dispensations in previous year

**Table S8.** Hazard ratios (95% CIs) for hospitalization with Covid-19 for all variables included as covariates in the Cox multivariable model with inverse probability weighting by the propensity score on population without smoking-related diseases.

| Characteristic<sup>†</sup> | HR* (95% CI) |
| --- | --- |
| Nicotine Replacement Therapy | 0.49 (0.40 – 0.60) |
| Deprivation Index |  |
| 1 (the least deprivation) | Ref |
| 2 | 1.17 (0.81 – 1.69) |
| 3 | 1.47 (1.02 – 2.11) |
| 4 | 1.60 (1.08 – 2.37) |
| 5 (the most deprivation) | 1.30 (0.89 – 1.89) |
| Unknown | 1.50 (0.44 – 5.10) |
| **Comorbidities** |  |
| Obesity | 1.63 (1.57 – 1.69) |
| Diabetes | 0.92 (0.90 – 0.95) |
| Dementia | 0.51 (0.41 – 0.62) |
| Alcohol use disorders | 3.38 (3.28 – 3.47) |
| Substance abuse | 2.82 (2.71 – 2.93) |
| Suicide attempt | 1.26 (1.20 – 1.33) |
| **Medications** |  |
| Anti-hypertensive drugs<sup>a</sup> |  |
| No | Ref |
| ACE | 1.14 (1.12 – 1.17) |
| Others | 0.99 (0.97 – 1.00) |
| Lipid lowering drugs<sup>a</sup> | 1.70 (1.67 – 1.73) |
| Antidepressants<sup>a</sup> | 1.58 (1.56 – 1.61) |
| Anxiolytics<sup>a</sup> | 1.87 (1.84 – 1.90) |
| Hypnotics<sup>a</sup> | 1.47 (1.43 – 1.52) |
| Neuroleptics<sup>a</sup> | 2.02 (1.97 – 2.08) |
| NSAID<sup>a</sup> | 1.49 (1.46 – 1.51) |
| Oral Corticosteroids<sup>a</sup> | 1.62 (1.56 – 1.67) |

† ACE Angiotensin-converting enzyme; CMU\_C complementary universal health insurance; NSAID Non-steroidal anti-inflammatory drug
§Paris and great Paris
<sup>a</sup>At least three dispensations in previous year
\*This Cox multivariable model, which was inverse probability weighted by the propensity score, was designed to control for potential confounding-by-indication of the exposure to nicotine-replacement therapy and hospitalization with Covid-19. It was additionally stratified according to sex, age, department and CMU-c, which parameter estimates are not generated or shown. The results in this table are provided for the reader's information but should not be interpreted to provide information on predictors or causes of the composite outcome

**Table S9.** Hazard ratios (95% CIs) for hospitalization with Covid-19 for all variables included as covariates in the Cox multivariable model with inverse probability weighting by the propensity score on population with exposed individual with smoking-related diseases.

| Characteristic<sup>†</sup> | HR* (95% CI) |
| --- | --- |
| Nicotine Replacement Therapy | 0.93 (0.75 – 1.16) |
| Deprivation Index |  |
| 1 (the least deprivation) | Ref |
| 2 | 1.26 (0.81 – 1.95) |
| 3 | 1.06 (0.67 – 1.67) |
| 4 | 0.86 (0.53 – 1.39) |
| 5 (the most deprivation) | 1.01 (0.65 – 1.59) |
| Unknown | 2.06 (0.67 – 6.34) |
| **Comorbidities** |  |
| Obesity | 1.60 (1.14 – 2.23) |
| Diabetes | 2.11 (1.47 – 3.05) |
| Dementia | 3.57 (0.67 – 18.88) |
| Alcohol use disorders | 1.76 (1.08 – 2.88) |
| Substance abuse | 0.95 (0.41 – 2.17) |
| Suicide attempt | 2.50 (0.81 – 7.73) |
| **Medications** |  |
| Anti-hypertensive drugs<sup>a</sup> |  |
| No | Ref |
| ACE | 1.08 (0.70 – 1.64) |
| Others | 1.06 (0.77 – 1.45) |
| Lipid lowering drugs<sup>a</sup> | 0.77 (0.56 – 1.05) |
| Antidepressants<sup>a</sup> | 1.27 (0.86 – 1.88) |
| Anxiolytics<sup>a</sup> | 1.07 (0.72 – 1.60) |
| Hypnotics<sup>a</sup> | 1.43 (0.84 – 2.42) |
| Neuroleptics<sup>a</sup> | 1.34 (0.75 – 2.40) |
| NSAID<sup>a</sup> | 1.74 (1.17 – 2.57) |
| Oral Corticosteroids<sup>a</sup> | 2.82 (1.83 – 4.35) |

<sup>†</sup> ACE Angiotensin-converting enzyme; CMU\_C complementary universal health insurance; NSAID Non-steroidal anti-inflammatory drug
<sup>§</sup>Paris and great Paris
<sup>a</sup>At least three dispensations in previous year
\*This Cox multivariable model, which was inverse probability weighted by the propensity score, was designed to control for potential confounding-by-indication of the exposure to nicotine-replacement therapy and hospitalization with Covid-19. It was additionally stratified according to sex, age, department and CMU-c, which parameter estimates are not generated or shown. The results in this table are provided for the reader's information but should not be interpreted to provide information on predictors or causes of the composite outcome

#### Figure

**Figure S1.**
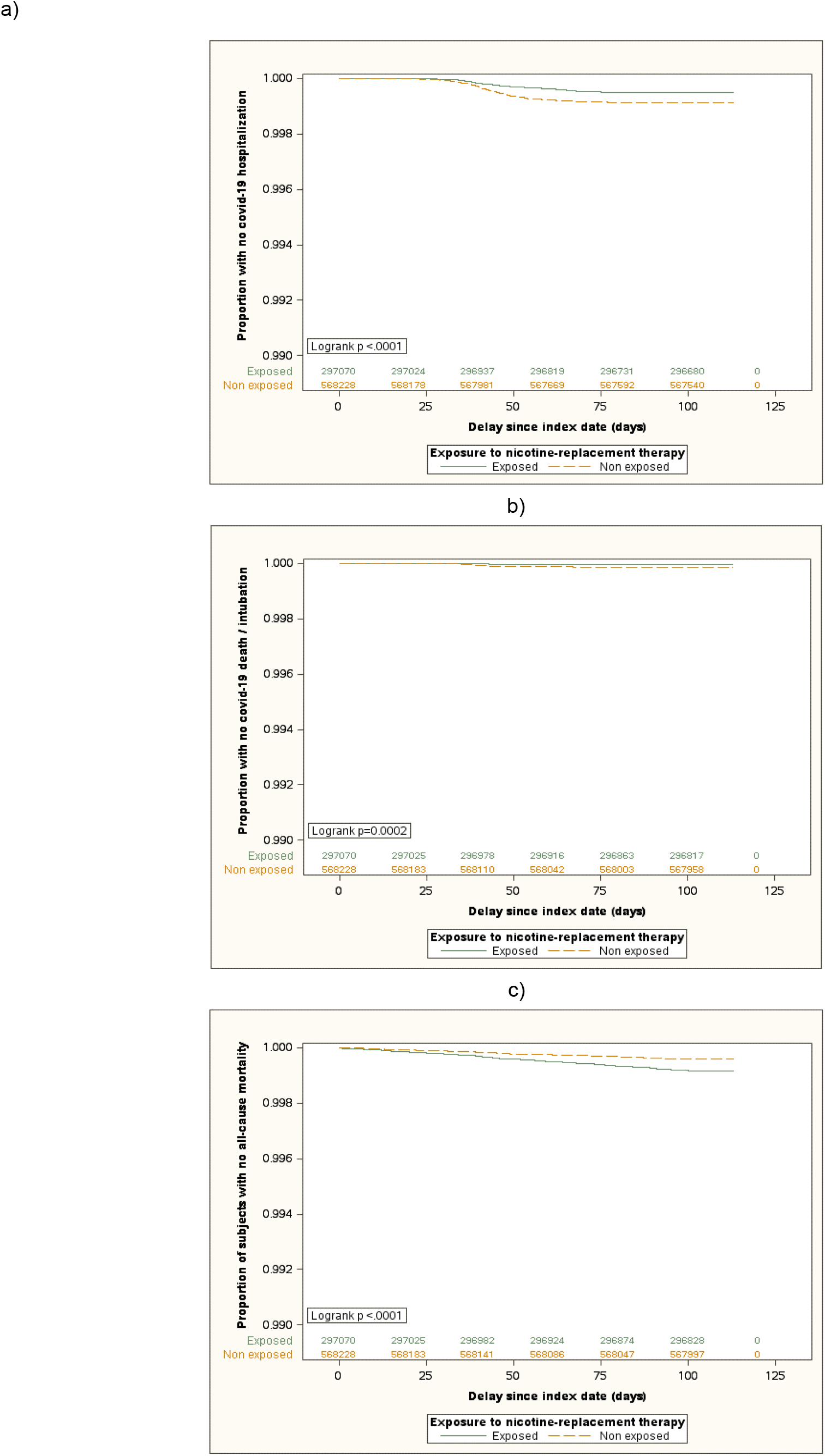
Kaplan-Meier Survival Curves of time to a) hospitalization for covid-19, b) intubation or death for covid-19 and c) all-cause mortality stratified by exposure group and showing the proportion of subject with no event from the index date (15th of February 2020) to the end of the follow-up on population without smoking-related diseases

**Figure S2.**
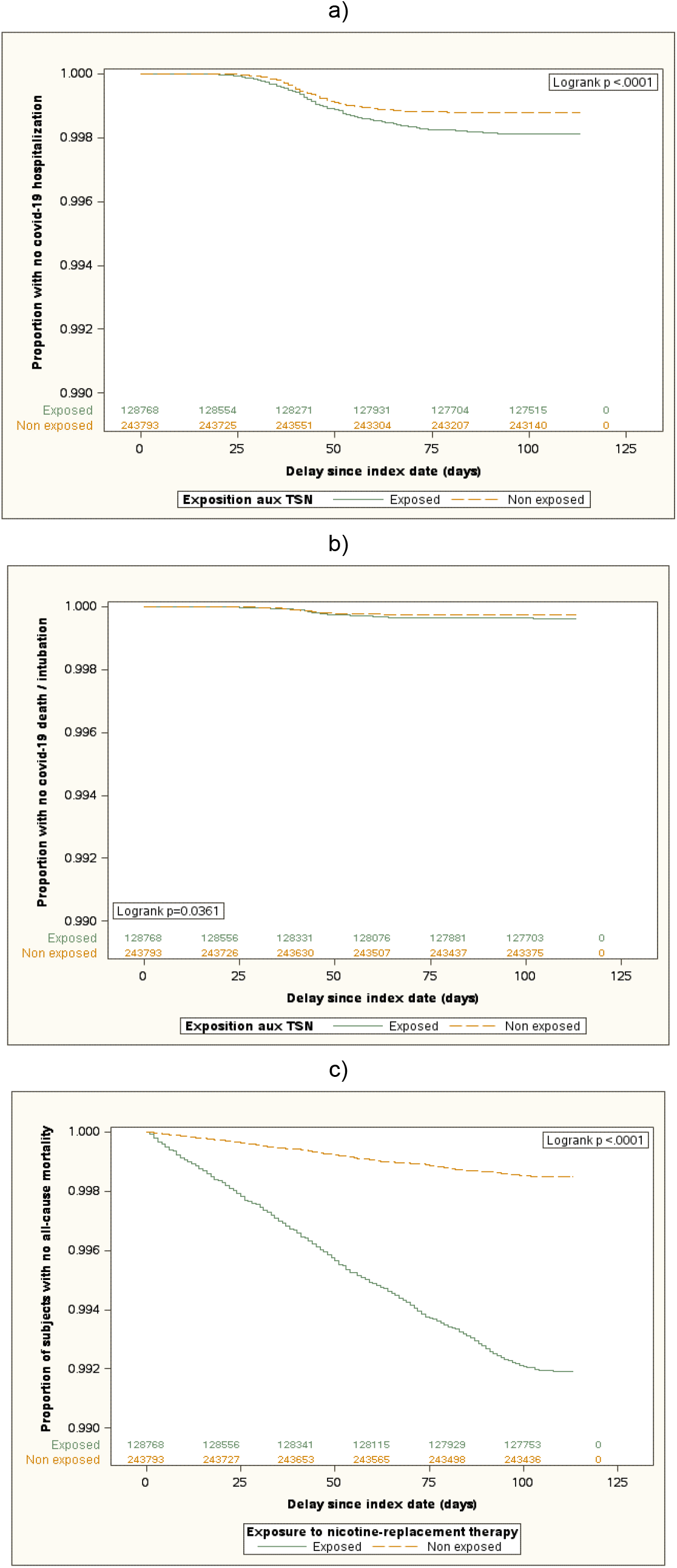
Kaplan-Meier Survival Curves of time to a) hospitalization for covid-19, b) intubation or death for covid-19 and c) all-cause mortality stratified by exposure group and showing the proportion of subject with no event from the index date (15th of February 2020) to the end of the follow-up on population with exposed individual with smoking-related diseases

